# Methylome-wide association study of early life stressors and adult mental health reveals a relationship between birth date and cell type composition in blood

**DOI:** 10.1101/2021.03.10.21253201

**Authors:** David M. Howard, Oliver Pain, Ryan Arathimos, Miruna C. Barbu, Carmen Amador, Rosie M. Walker, Bradley Jermy, Mark J. Adams, Ian J. Deary, David Porteous, Archie Campbell, Patrick F. Sullivan, Kathryn L. Evans, Louise Arseneault, Naomi R. Wray, Michael Meaney, Andrew M. McIntosh, Cathryn M. Lewis

**Author notes:** Corresponding author: David M. Howard, Social, Genetic and Developmental Psychiatry Centre, Institute of Psychiatry, Psychology & Neuroscience, King’s College London, UK.

## Abstract

The environment and events that we are exposed to in utero, during birth and in early childhood influence our future physical and mental health. The underlying mechanisms that lead to these outcomes in adulthood are unclear, but long-term changes in epigenetic marks, such as DNA methylation, could act as a mediating factor or biomarker. DNA methylation data was assayed at 713,522 CpG sites from 9,537 participants of the Generation Scotland: Scottish Family Health Study, a family-based cohort with extensive data on genetic, medical, family history and lifestyle information. Methylome-wide association studies of eight early life environment phenotypes and two adult mental health phenotypes were conducted using DNA methylation data collected from adult whole blood samples. Two genes involved with different developmental pathways (PRICKLE2 and ABI1) were annotated to CpG sites associated with preterm birth (P < 1.27 × 10 ^−9^). A further two genes important to the development of sensory pathways (SOBP and RPGRIP1) were annotated to sites associated with low birth weight (P < 4.35 × 10^−8^). Genes and gene-sets annotated from associated CpGs sites and methylation profile scores were then used to quantify any overlap between the early life environment and mental health traits. However, there was no evidence of any overlap after applying a correction for multiple testing. Time of year of birth was found to be associated with a significant difference in estimated lymphocyte and neutrophil counts. Early life environments influence the risk of developing mental health disorders later in life; however, this study provides no evidence that this is mediated by stable changes to the methylome detectable in peripheral blood.

## Introduction

The diathesis-stress model posits that behaviours and psychological disorders are the result of underlying biological factors (diatheses) plus exposure to stressful events or environments. Childhood adversity increases the risk of poorer physical and mental health outcomes in later life [1, 2], with neglect, sexual and emotional abuse, and violence providing greater risk [3-6]. However, multiple studies have reported additional perinatal and early life stressors and environments that also impact on adult mental health. Individuals born preterm or with a low birth weight are more likely to experience problems with attentiveness and hyperactivity, as well as elevated levels of anxiety and depression [7-9]. The time of year of birth has also been shown to influence psychiatric outcomes in later life [10-12], with those born in January having higher risk of schizophrenia and bipolar and those born in June and July having greater risk of depression. Parental factors, including the age of the parents at birth [13] or having an absent parent [14], are reported to increase the risk of depression. Where we live has also been shown to be detrimental to mental health [15] and is likely due to a variety of environmental and social factors [16, 17].

Adverse environments in the gestational and postnatal periods are known to cause long-term alterations to DNA methylation across the genome [18]. These modifications are reported to mediate resilience and responses to stress-related disorders throughout the life course [19]. Epigenomic variation has also been implicated in a range of psychiatric conditions [20], including bipolar disorder [21], schizophrenia [22], and major depressive disorder (MDD) [23]. Much of the epigenetic research seeking to link early life environments and MDD has been performed using small cohort samples sizes (typically less than 1,000) and focussing on candidate regions [24]. However, this approach is likely to be suboptimal as demonstrated by the lack of reproducible results from candidate gene association studies for MDD [25].

The principal aim of the current research was to investigate whether alterations to DNA methylation have the potential to mediate the stress component in the diathesis-stress model. This was achieved by conducting a methylome-wide association study (MWAS) of early life environments and later mental health outcomes in a single study cohort of over 9,500 adults. Linked electronic health records, responses at interview and questionnaire data were used to ascertain early life environments, with MDD and brief resilience scale (BRS) assessed as the mental health outcomes. The associated CpG sites identified by the MWAS of these phenotypes were then annotated to genes and gene-sets. The gene and gene-set overlap between the early life environments and mental health outcomes were then examined. Additionally, methylation profile scores were used to assess the broader methylome-wide overlap between the early life environment and mental health traits. The influence of time of year of birth on estimated blood cell type composition was investigated further in an independent study cohort.

## Materials & Methods

### Generation Scotland: Scottish Family Health Study (GS:SFHS)

GS:SFHS [26] is a family-based cohort study of 24,080 participants (14,154 female and 9,926 male) aged between 18 and 100 (mean = 47.6 years, standard deviation = 15.4 years). Baseline data was collected between 2006 and 2011 and covered medical, behaviour and lifestyle factors with a subset recontacted (N = 9,618) in 2015 and 2016 with additional phenotypes collected [27]. Clinical information from linked electronic hospital records across the life course was also available. At the baseline appointment, a blood draw was taken from each participant, which has so far been used to obtain DNA methylation data for 9,773 individuals.

### Phenotypes

Multiple phenotypes were generated from the baseline and recontact data focused on either early life environments or adult mental health. The early life environments were broadly categorised as either biological (preterm birth, low birth weight, and time of year of birth) or sociodemographic (having a lone parent, having a young parent, urbanicity, and population density). The adult mental health measures were MDD and psychological resilience (measured using the BRS). The demographics of GS:SFHS are provided in Table 1.

**Table 1.** Demographic information for Generation Scotland: Scottish Family Health Study

| Item | Whole sample |  |  | Methylation sample |  |  |
| --- | --- | --- | --- | --- | --- | --- |
|  | N | Present / absent (%) | Mean, St. dev. | N | Present / absent (%) | Mean, St. dev. |
| Female | 24,080 | 14,154 / 9,926 (58.8) |  | 9,537 | 5,638 / 3,899 (59.1) |  |
| Age | 24,080 |  | 47.64, 15.41 | 9,537 |  | 49.81, 13.70 |
| Preterm birth | 3,134 | 109 / 3,025 (3.5) |  | 950 | 31 / 919 (3.3) |  |
| Low birth weight | 3,129 | 103 / 3,026 (3.3) |  | 949 | 30 / 919 (3.2) |  |
| Birth month | 24,080 | 12,842 / 11,238 (53.3) |  | 9,516 | 5,459 / 4,057 (57.4) |  |
| Birth date | 22,656 |  | 0.01, 0.71 | 9,509 |  | 0.01, 0.71 |
| Young parent | 23,546 | 1,241 / 22,305 (5.3) |  | 9,398 | 534 / 8,864 (5.7) |  |
| Lone parent | 22,543 | 1,333 / 21,210 (5.9) |  | 9,086 | 580 / 8,506 (6.4) |  |
| Urban environment | 19,428 | 11,739 / 7,689 (60.4) |  | 8,017 | 5,468 / 2,549 (68.2) |  |
| Population density | 19,426 |  | 1,949.72, 1,611.32 | 8,017 |  | 2019.44, 1,489.35 |
| Major depressive disorder | 21,340 | 2,766 / 18,574 (13.0) |  | 9,481 | 1,624 / 7,857 (17.1) |  |
| Brief resilience scale | 9,354 |  | 0.00, 3.80 | 4,839 |  | -0.19, 3.90 |

Number of individuals (N) available for each item. The present N, absent N and prevalence (%) are reported for binary items with mean and standard deviation (St. dev.) reported for continuous items.

Preterm births were categorized as a recorded gestation of period of less than 37 weeks (based on World Health Organization guidance [28]) using the SMR02 - Maternity Inpatient and Day Case linked electronic health records. These records were available for individuals born after 1992. Birth weight for GS:SFHS participants was also obtained from SMR02 electronic health records. The threshold for low birth weight was based on the sex and gestation length adjusted 3^rd^ centile for birth weight reported using a Scottish sample and SMR02 records by Bonellie et al. [29].

The time of year of birth was assessed using two phenotypes. Firstly, a binary phenotype for birth month was generated with those born between April and October inclusive compared to those born during the remaining months. These months were selected based on the review of birth month and depression by Schnittker [12], with increased risk of depression reported for those born April through October. Secondly, the birth date during the year was assessed as a continuous phenotype (*y*) and calculated as:

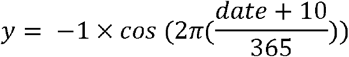

Where *y* ranged from −1 for those born on the winter solstice (21^st^ December) to +1 for those born on the summer solstice (21^st^ June). *date* was the day of birth during the year for each participant (January 1^st^ = 1, January 2^nd^ = 2, etc.). For those born on the 29^th^ February, *date* = 59.5.

Using the self-reported age of an individual’s parents at the time of birth, an individual was classified as having a young parent if either parent was under the age of 21. Where an individual was recorded as having a lone parent (see below), the age of the parent that the individual lived with was used.

An individual was recorded as having a lone parent if either their mother or father was recorded as living in a different country or region to the individual and the other parent at the time of birth of the individual. The regions reflect the 32 council areas of Scotland. Individuals with missing information, or where an individual was reported not to live with either parent, were excluded.

The region that an individual was living in at the time of their birth was also used to define an urban environment phenotype. Individuals living in Edinburgh, Glasgow, Aberdeen, or Dundee were classified as being urban and those living in other regions as non-urban. The year of an individual’s birth and the region they were living in were also used to obtain a measure of population density based on population estimates from the National Records of Scotland [30] and was recorded as the number of individuals per square kilometre.

To determine MDD status, the initial screening questions from the Structured Clinical Interview for DSM-IV (SCID) Non-Patient Version [31] were used to identify those individuals that would subsequently complete the mood sections of the SCID. The SCID was administered by nurses trained in its application and further information on the MDD criteria used in GS:SFHS is reported in Fernandez-Pujals et al. [32]. Participants who met the criteria for at least one MDD episode in the mood sections of the SCID were classified as cases and those who did not meet this criteria or did not report MDD symptoms in the initial screening were classified as controls.

The BRS [33] was used to determine a continuous measure of an individual’s psychological resilience and ability to ‘bounce back’ from stressful events. BRS was obtained for a subset (N = 9,505) of the cohort during a recontact of all GS:SFHS participants in 2015 and 2016. BRS was ascertained from the response to six questions using a 5-point Likert scale with further details on the assessment provided by Navrady et al. [27]. Individuals that responded to 5 or more questions were retained (N = 9,354) with missing values imputed using the missForest package [34] applying 500 trees per forest. A principal component analysis using the Psych package [35] was applied to the data to extract the first unrotated principal component which was then scaled to create a continuous measure of BRS.

### Regression of adult mental health on early life environments

To determine whether there was an association between early life stressors and the measures of adult mental health a generalized linear mixed model was used from the lme4qtl package [36]. MDD and BRS were assessed separately as the dependent variable for an association with each early life factor in turn as the independent variable. The total genetic value derived from pedigree data (with the variance/covariance structure defined by a kinship matrix using the kinship2 package [37]) was fitted as a random effect, jointly with sex fitted as a fixed effect. Binary traits were fitted as factors and continuous traits were centered and scaled to have a mean of 0 and a standard deviation of 1. A binomial regression with a logit function was used to assess associations with MDD. For a significant association between a mental health trait and an early life environment a Bonferroni correction was applied to the P-values within each mental health trait, P < 6.25 × 10^−3^ (α = 0.05 / 8).

### Methylation Data

The Infinium MethylationEPIC BeadChip (Illumina Inc.) was used to profile DNA methylation data at 853,307 CpG sites generated in two sets of individuals. There were 5,190 individuals in Set 1 and 4,583 unrelated individuals in Set 2, with no related individuals between the sets. Quality control was then applied to both Sets and full details are provided by Barbu et al. [38] and McCartney et al. [39]. In summary, individuals were excluded if they were outliers based on multi-dimensional scaling or the predicted sex from the methylation data mismatched the recorded sex. CpG sites were excluded based on low beadcount, poor detection P-value, or if they were predicted to bind sub-optimally. The log2 ratio of the intensities of methylated probe versus unmethylated probe data was used to generate methylation M-values separately in each Set [40]. In total there were 713,522 CpG sites remaining with 5,087 individuals in Set 1 and 4,450 individuals in Set 2.

After combining the Sets (9,537 individuals), correction was applied for:

1. technical variation, where M-values were included as dependent variables in a mixed linear model adjusting for the plate used to profile the DNA methylation data and the date of the individual’s blood draw as random effects, jointly with plate position, Set, clinic, year of appointment, day of week of appointment, and the first 10 principal components (from the EPIC array control sites) as fixed effects; and
2. biological variation by fitting residuals of (1) as dependent variables in a second mixed linear model adjusting for genetic and common family shared environmental contributions (classed as G: common genetic; K: kinship; F: nuclear family; C: couple; and S: sibling, see Xia et al. [41] and Zeng et al. [42] for further information) as random effects, jointly with sex, age, and estimated blood cell type composition (CD8T, CD4T, NK, Bcell, Mono, Gran; obtained using the Houseman algorithm [43] within the Meffil package [44]) as fixed effects.

### Methylome-wide association study of early life environments and adult mental health

Two methods (MWAS 1 and MWAS 2) were used to conduct the methylome-wide association study generating two sets of summary statistics for each phenotype. MWAS 1 was conducted using the eBayes function within the limma package [45], which applies a linear mixed model to the data. The M-values, corrected for technical and biological variation, were the dependent variable with each phenotype included separately as the independent variable. Set, smoking status (ever or never), smoking pack years, and the first 20 principal components created from the M-values using the FactoMineR package [46], were fitted as fixed effects.

MWAS 2 was conducted using the software tool OmicS-data-based Complex trait Analysis (OSCA) [47]. First, to account for methylome-wide correlational structure, this tool uses a linear regression analysis to identify groups of lead CpG sites based on the association test statistics. Each phenotype in turn was the dependent variable with the corrected M-values as the independent variable adjusted for the same covariates used in MWAS 1 (Set, smoking status, smoking pack years, and the first 20 principal components). Second, each group of lead sites were fitted as random effects in a Multi-component MLM-based association excluding the target (MOMENT) analysis to assess the effect of each target probe in turn, with the respective phenotype as the dependent variable and the M-values as the independent variable. MOMENT also fits predicted blood cell type composition (basophil, eosinophil, lymphocyte, monocyte, and neutrophil; based on haematological analysis of the Lothian Birth Cohorts 1921 and 1936 (LBC) [48]) as a fixed effect.

A methylome-wide significance threshold was determined using a Bonferroni correction based on the number of CpG sites analysed for each analysis: P < 7.01 × 10^−8^ (α = 0.05 / 713,522). To visualise the MWAS output, Miami plots were created using ggplot2 [49] with the P-values for MWAS 1 on the log_10_ scale and for MWAS 2 on the -log_10_ scale. QQ-plots were created using Haplin [50] with the shaded error representing the 95% confidence interval. Genomic inflation (λ) was calculated for each output as the median of the observed chi-squared distribution of P-values divided by the median of the expected chi-squared distribution.

### Gene and Gene set analysis

The significant CpG sites (P < 7.01 × 10^−8^) for each phenotype and each MWAS were annotated to genes based on location. For MWAS 1, the missMethyl package [51] was used to annotate genes according to location. For MWAS 2, the annotations reported by the OSCA package were used. The gometh function within missMethyl was used to analyse the data for enrichment of Gene Ontology and KEGG gene sets. The overlap of the annotated genes and enriched gene sets between the early life environment and adult mental health phenotypes were examined.

### Methylation profile scores

MWAS 2 was used to calculate summary statistics for each early life environment phenotype using the individuals in Set 1 (N = 5,190). Multiple P-value thresholds (< 10^−7^, < 10^−6^, < 10^−5^, < 10^−4^, < 10^−3^, < 10^−2^, < 10^−1^) were used to identify those sites for inclusion. For each individual in Set 2, an aggregated methylation profile score was calculated by multiplying the CpG effect sizes from Set 1 by their respective M-values in Set 2. There were no related individuals between Set 1 and Set 2. The ability of these profile scores to predict the same phenotype, MDD or BRS, was assessed after adjusting for sex, age, and the first 20 principal components (derived from single nucleotide polymorphism data) as fixed effects. For prediction of binary phenotypes, a coefficient of determination was calculated using Nagelkerke’s R^2^ [52] using a population prevalence of that equal to the sample. For quantitative phenotypes, a simple linear regression was used with the adjusted R^2^ reported. A Bonferroni correction was used to identify significant prediction based on the number of early life phenotypes examined in each case: *P* < 6.25 × 10^−3^ (α = 0.05 / 8).

### Blood cell type composition

A notable difference was observed between the results from MWAS 1 and MWAS 2 for the time of year of birth phenotypes. The inclusion of predicted cell type composition as a fixed effect in MWAS 2 represents a potential source of these differences. To investigate cell type composition further, haematological and DNA methylation data from 1,308 participants of the LBC was obtained [48, 53], mirroring the development of the OSCA package used in MWAS 2. A similar protocol to that described by Zhang et al. [47] was used to identify effect sizes and *P*-values between 214,923 CpG sites and the cell counts of basophils, eosinophils, lymphocytes, monocytes, and neutrophils. First, the significant lead CpG sites (*P* < 7.01 × 10^−8^) identified for birth date for MWAS 1 in GS:SFHS were examined in the LBC for an association with the five cell type compositions using QQ-plots and λ. Lead sites were identified by retaining the site with the lowest P-value amongst sites with an r^2^ greater than 0.1 within a 1 Mb window. Second, methylation profile scores were created for individuals in the LBC using the effect sizes from MWAS 1 for birth date in GS:SFHS. These profile scores were created using lead CpG sites at seven P-values thresholds (< 10^−7^, < 10^−6^, < 10^−5^, < 10^−4^, < 10^−3^, < 10^−2^, < 10^−1^). A simple linear regression was then used to calculate the association between the profile scores for birth date and the five measured cell type compositions, after adjusting for the sex and age of participants as fixed effects. A Bonferroni correction based on the five of cell types analysed required a P-value < 0.01 (α = 0.05 / 5) for an association.

## Results

We conducted analyses of early life environments and adult mental health using the Generation Scotland: Scottish Family Health Study cohort (n = 24,080). First, we used regression to assess the association between early life environments and MDD and BRS using phenotypic data. Second, we conducted methylome-wide association analysis to identify CpG sites associated with these phenotypes. Finally, we used the summary statistics from the methylome-wide association analyses to determine the extent of any overlap between phenotypes.

### Regression of adult mental health on early life environments

Generalised linear mixed models were used to assess each early life environment in turn and its association with either MDD or BRS (Supplementary Table 1), after adjusting for sex and relatedness between individuals. There were no significant associations between the early life environments and the adult mental health phenotypes after adjusting for multiple testing (*P* > 6.25 × 10^−3^). All early life environments, except for preterm birth, marginally increased the risk of developing MDD in adulthood. All early life environments, except for having a lone parent, marginally lowered BRS suggesting a negative effect on psychological resilience. The associations between time of year of birth and BRS were nominally significant (P < 0.05) with those born across the summer months scoring lower on the BRS; however, these associations did not remain after correction for multiple testing.

### Methylome-wide association study of early life environments and adult mental health

Normalised M-values for 713,522 CpG sites profiled from combined blood samples of 9,537 GS:SFHS individuals (5,087 in Set 1 and 4,450 in Set 2) remained after quality control procedures. The associations between these M-values and the early life environments and mental health phenotypes were calculated using two association study methods (MWAS 1 and MWAS 2). MWAS 1 fits a linear regression model to the data with the M-values as the dependent variable, whereas MWAS 2 fits the phenotype as the dependent variable while accounting for methylome-wide correlational structure and with an additional correction for predicted blood cell type composition.

The significant CpG sites (*P* < 7.01 × 10^−8^) from MWAS 1 and MWAS 2 are in Table 2, except those associated with birth month of birth date. The six significant CpG sites identified in MWAS 2 were also significant in MWAS 1 for the same phenotypes. Supplementary Table 2 and Supplementary Table 3 contains the significant sites (*P* < 7.01 × 10^−8^) for the birth month and birth date phenotypes, respectively. In MWAS 1, there were 93 significant sites for birth month and 637 significant sites for birth date. However, no CpG sites were significant in MWAS 2 for either birth month or birth date with further analysis covered in the blood cell type composition section.

**Table 2.** CpG sites associated with phenotypes in either MWAS 1 or MWAS 2

| Phenotype | CpG site | Chr | BP position | Annotated Gene | MWAS 1 |  |  | MWAS 2 |  |  |
| --- | --- | --- | --- | --- | --- | --- | --- | --- | --- | --- |
|  |  |  |  |  | Effect size | Standard error | P-value | Effect size | Standard error | P-value |
| Preterm birth | cg00725333 | 3 | 64189256 | *PRICKLE2* | -0.2747 | 0.0376 | **$6.24 \times 10^{-13}$** | -0.0364 | 0.0060 | **$1.15 \times 10^{-9}$** |
|  | cg17668848 | 3 | 108029973 | *HHLA2* | -0.2871 | 0.0396 | **$9.13 \times 10^{-13}$** | -0.0356 | 0.0058 | **$8.21 \times 10^{-10}$** |
|  | cg24329141 | 10 | 27095369 | *ABI1* | -0.2368 | 0.0352 | **$2.98 \times 10^{-11}$** | -0.0345 | 0.0057 | **$1.27 \times 10^{-9}$** |
| Low birth weight | cg15582176 | 1 | 1183528 | *C1QTNF12* | -0.3604 | 0.0661 | **$6.53 \times 10^{-8}$** | -0.0254 | 0.0054 | $2.49 \times 10^{-6}$ |
|  | cg19909717 | 6 | 107924415 | *SOBP* | -0.2225 | 0.0403 | **$4.35 \times 10^{-8}$** | -0.0329 | 0.0055 | **$1.59 \times 10^{-9}$** |
| | cg21803443 | 7 | 126547786 | *GRM8* | -0.3656 | 0.0618 | **$4.62 \times 10^{-9}$** | -0.0278 | 0.0056 | $7.22 \times 10^{-7}$ |
|  | cg12090821 | 14 | 21755658 | *RPGRIP1* | -0.0857 | 0.0152 | **$2.43 \times 10^{-8}$** | -0.0329 | 0.0058 | **$1.47 \times 10^{-8}$** |
| | cg05905731 | 16 | 85485785 | | -0.1695 | 0.0300 | **$2.23 \times 10^{-8}$** | -0.0294 | 0.0055 | $7.50 \times 10^{-8}$ |
| Young parent | cg00528572 | 11 | 92703433 | *MTNR1B* | -0.1369 | 0.0200 | **$9.04 \times 10^{-12}$** | -0.0145 | 0.0025 | **$4.47 \times 10^{-9}$** |
| | cg02427109 | 18 | 77917459 | *PARD6G, PARD6G-AS1* | -0.1163 | 0.0213 | **$4.96 \times 10^{-8}$** | -0.0101 | 0.0025 | $4.12 \times 10^{-5}$ |
| Population density | cg06759845 | 1 | 156460474 | *MEF2D* | $5.20 \times 10^{-6}$ | $9.40 \times 10^{-7}$ | **$3.40 \times 10^{-8}$** | 90.39 | 17.02 | $1.10 \times 10^{-7}$ |
| | cg12433043 | 6 | 100619893 | | $-6.65 \times 10^{-6}$ | $1.20 \times 10^{-6}$ | **$3.37 \times 10^{-8}$** | -88.86 | 16.75 | $1.13 \times 10^{-7}$ |
| | cg03623878 | 13 | 113655560 | *MCF2L* | $9.10 \times 10^{-6}$ | $1.58 \times 10^{-6}$ | **$9.56 \times 10^{-9}$** | 81.18 | 16.88 | $1.51 \times 10^{-6}$ |
| | cg08036492 | 17 | 13976536 | *COX10* | $3.09 \times 10^{-6}$ | $5.71 \times 10^{-7}$ | **$6.74 \times 10^{-8}$** | 87.75 | 18.29 | $1.61 \times 10^{-6}$ |
| MDD | cg02280719 | 1 | 6802222 | | 0.0400 | 0.0064 | **$5.99 \times 10^{-10}$** | 0.0174 | 0.0043 | $5.32 \times 10^{-5}$ |
| | cg08548783 | 6 | 57903690 | | 0.0378 | 0.0067 | **$1.37 \times 10^{-8}$** | 0.0142 | 0.0043 | $9.79 \times 10^{-4}$ |

MWAS 1 results are from a methylome-wide association analysis using linear regression and MWAS 2 results are from a methylome-wide association analysis using OmicS-data-based Complex trait Analysis. Bold P-values indicate an association after correction for multiple testing ( $P < 7.01 \times 10^{-8}$ ). Sites are ordered by phenotype and then genomic position. Chromosome number (Chr) and base pair (BP) position are based on genome assembly GRCh37 (hg19). Annotation of genes is provided by missMethyl. MDD = Major Depressive Disorder.

**Table 3.** Prediction of major depressive disorder and brief resilience scale in Set 2 using methylation profile scores. Scores were calculated using the CpG site effect sizes from methylome-wide association studies of early life environment phenotypes in Set 1

| Early life environment | Major depressive disorder |  |  | Brief resilience scale |  |  |
| --- | --- | --- | --- | --- | --- | --- |
|  | Threshold | R<sup>2</sup> | P-value | Threshold | R<sup>2</sup> | P-value |
| Preterm birth | $1 \times 10^{-4}$ | 0.0023 | 0.034 | $1 \times 10^{-7}$ | 0.0001 | 0.74 |
| Low birth weight | $1 \times 10^{-5}$ | 0.0005 | 0.32 | $1 \times 10^{-6}$ | 0.0009 | 0.21 |
| Birth month | $1 \times 10^{-6}$ | 0.0021 | 0.043 | $1 \times 10^{-6}$ | 0.0032 | 0.016 |
| Birth date | $1 \times 10^{-5}$ | 0.0013 | 0.10 | $1 \times 10^{-5}$ | 0.0016 | 0.09 |
| Young parent | $1 \times 10^{-3}$ | 0.0006 | 0.28 | $1 \times 10^{-5}$ | 0.0010 | 0.19 |
| Lone parent | $1 \times 10^{-5}$ | 0.0005 | 0.34 | $1 \times 10^{-4}$ | 0.0001 | 0.66 |
| Urban environment | $1 \times 10^{-5}$ | 0.0004 | 0.40 | $1 \times 10^{-2}$ | 0.0008 | 0.25 |
| Population density | $1 \times 10^{-4}$ | 0.0015 | 0.08 | $1 \times 10^{-6}$ | 0.0010 | 0.19 |

The effect sizes in Set 1 were obtained from methylome-wide association studies using OmicS-data-based Complex trait Analysis. Multiple thresholds based on CpG site $P$ -values were examined, with the threshold explaining the greatest phenotypic variance reported. Nagelkerke's $R^2$ was used to calculate the proportion of phenotypic variance explained on the liability scale for major depressive disorder. An adjusted $R^2$ from a simple linear regression was reported for brief resilience scale. The $P$ -value required for significance after correction for multiple testing was $P < 6.25 \times 10^{-3}$ .

Miami plots for preterm birth, low birth weight, having a young parent and MDD are in Figures 1-4, respectively. The remaining Miami plots are in Supplementary Figures 1-6 and QQ-plots and λ for all phenotypes are in Supplementary Figures 7-16.

**Figure 1.**
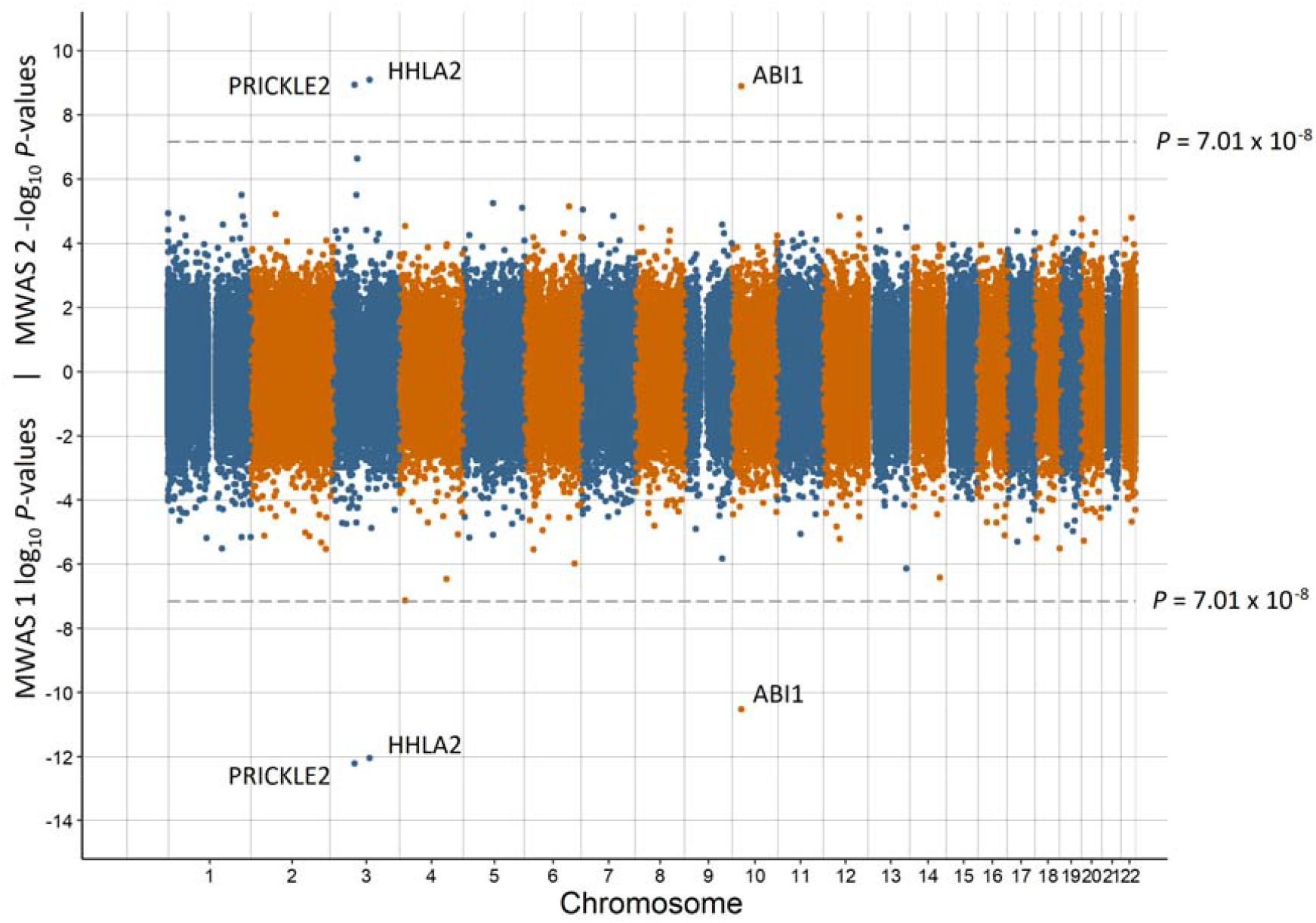
Miami plot of the observed P-values of each CpG site for an association with preterm birth Log_10_ P-values are shown for MWAS 1 and -log_10_ P-values are shown for MWAS 2. The dotted lines indicate methylome-wide significance (*p* = 7.01 × 10^−8^). The annotation of genes for significant sites is reported by missMethyl for MWAS 1 and by OSCA for MWAS 2

**Figure 2.**
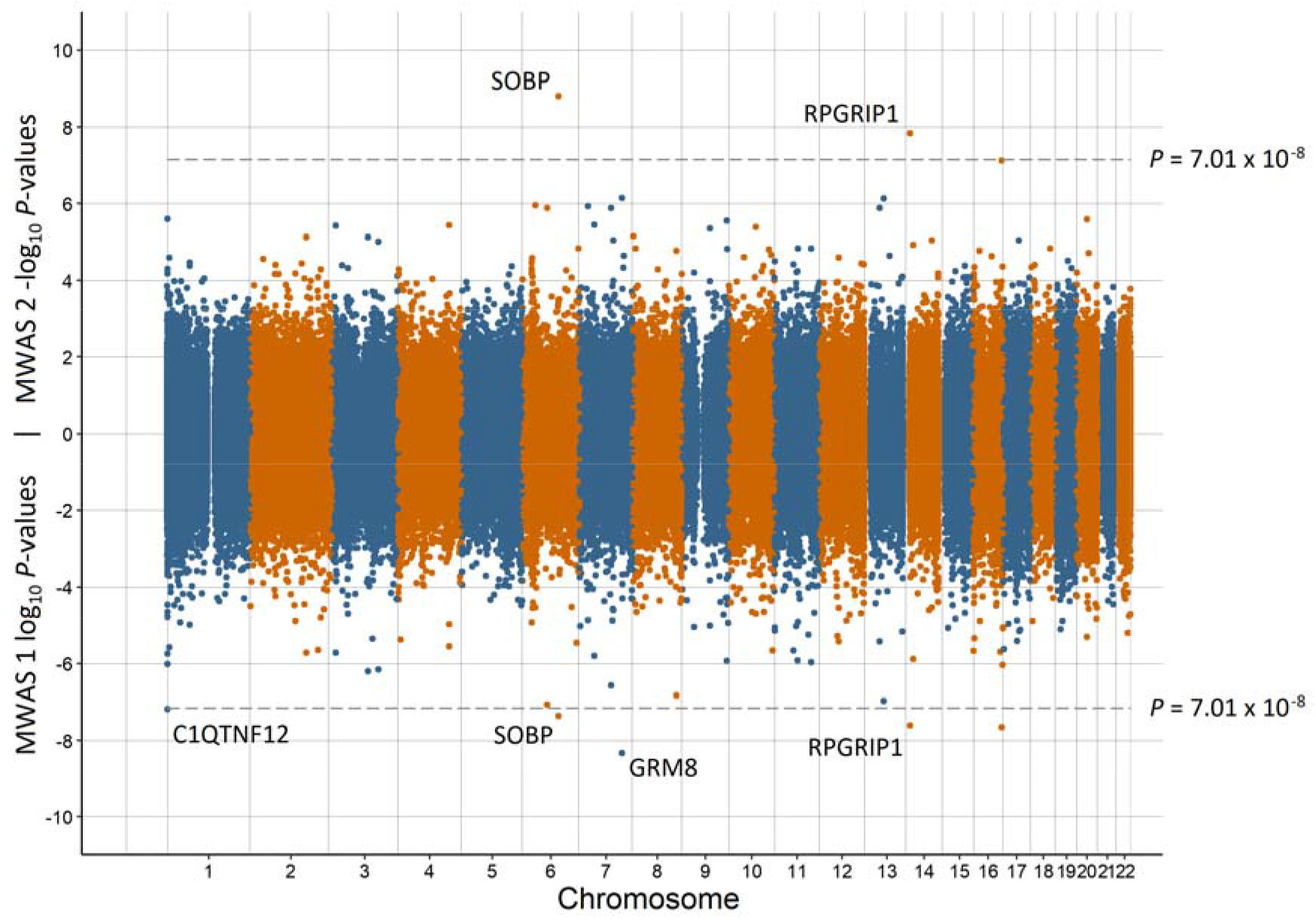
Miami plot of the observed P-values of each CpG site for an association with low birth weight Log_10_ P-values are shown for MWAS 1 and -log_10_ P-values are shown for MWAS 2. The dotted lines indicate methylome-wide significance (*p* = 7.01 × 10^−8^). The annotation of genes for significant sites is reported by missMethyl for MWAS 1 and by OSCA for MWAS 2

**Figure 3.**
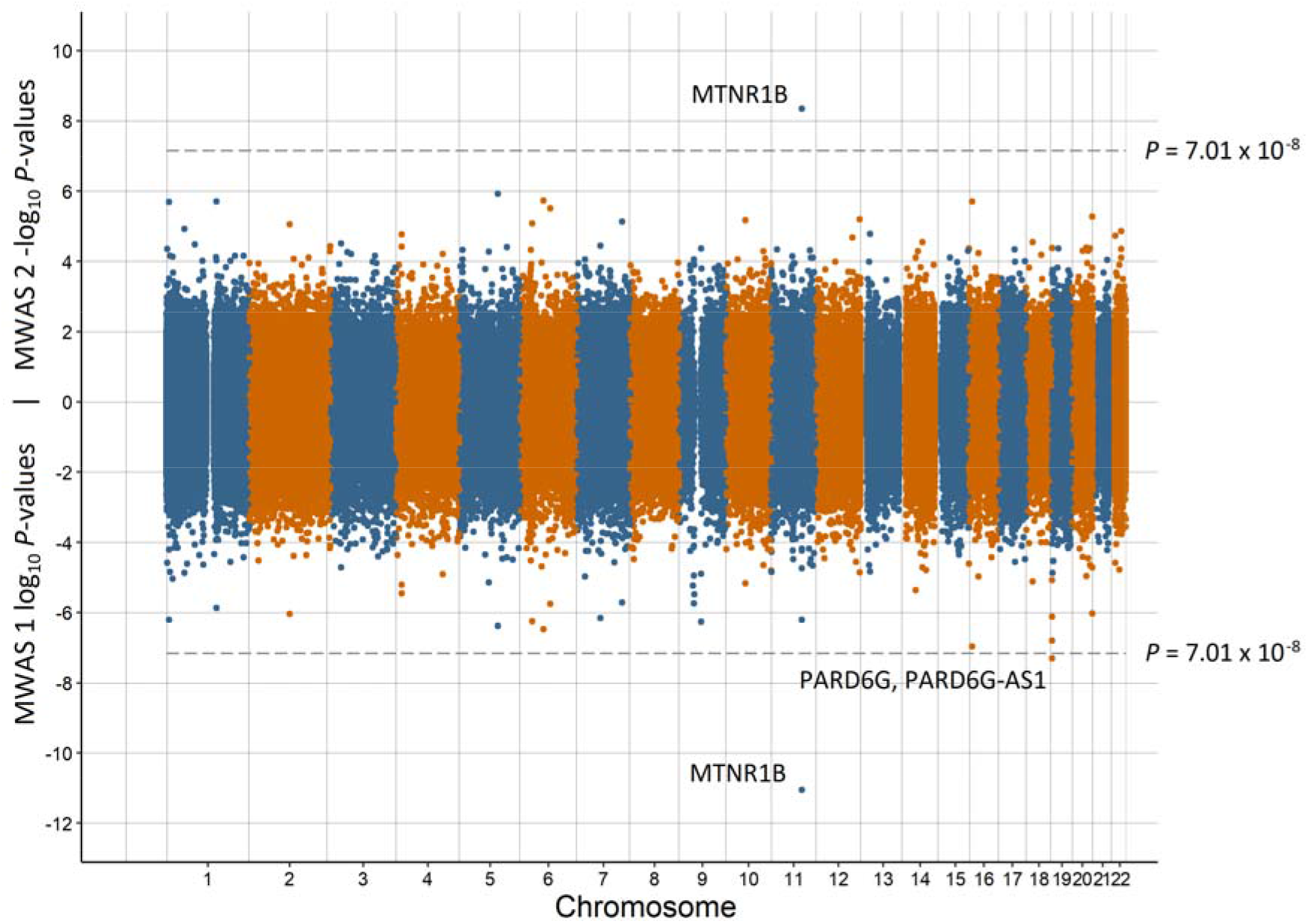
Miami plot of the observed P-values of each CpG site for an association with having a young parent Log_10_ P-values are shown for MWAS 1 and -log_10_ P-values are shown for MWAS 2. The dotted lines indicate methylome-wide significance (*p* = 7.01 × 10^−8^). The annotation of genes for significant sites is reported by missMethyl for MWAS 1 and by OSCA for MWAS 2

**Figure 4.**
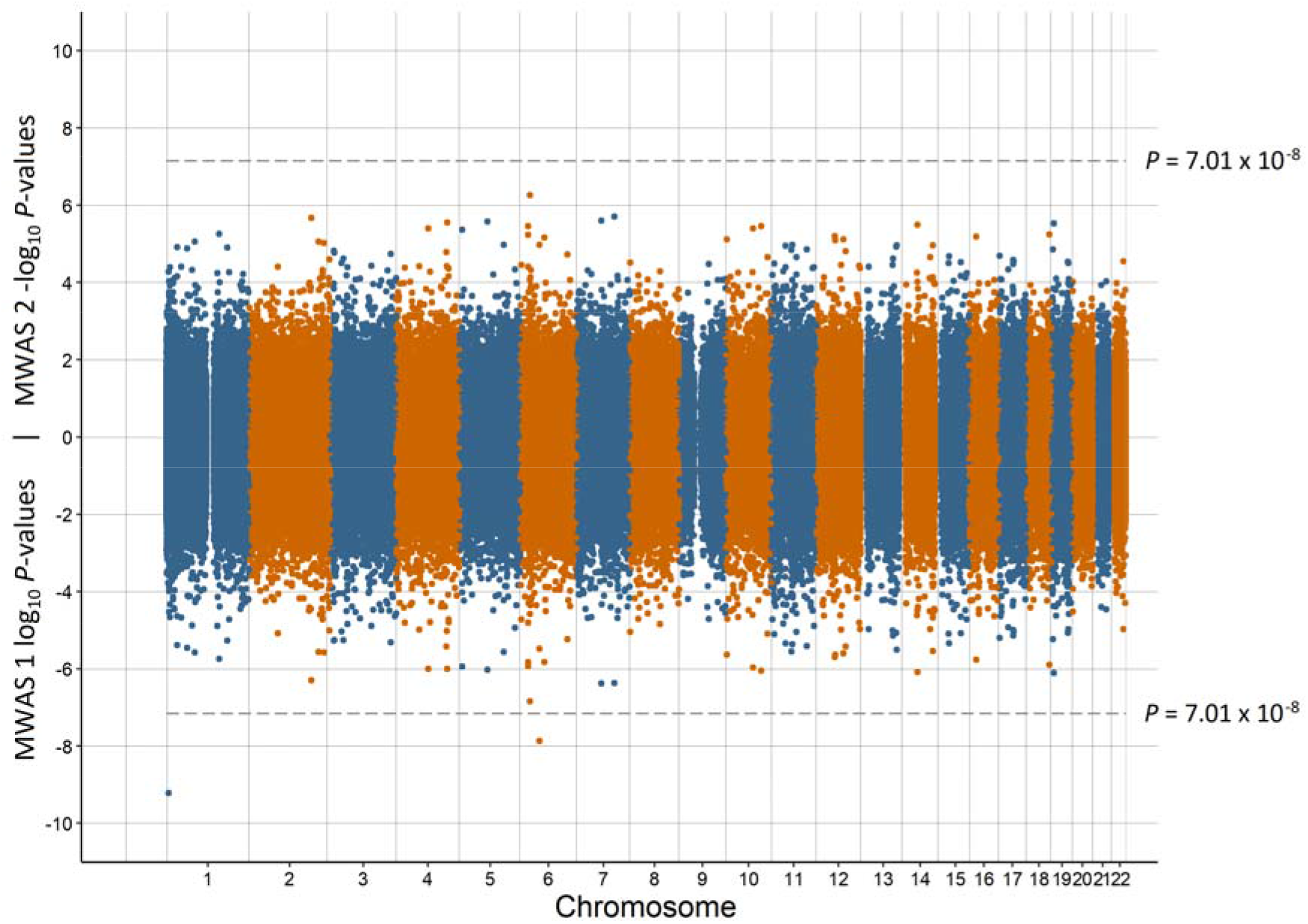
Miami plot of the observed P-values of each CpG site for an association with major depressive disorder Log_10_ *P*-values are shown for MWAS 1 and -log_10_ P-values are shown for MWAS 2. The dotted lines indicate methylome-wide significance (*p* = 7.01 × 10^−8^). Significant sites did not annotate to any genes.

### Genes and gene set analysis

The genes annotated to associated CpG sites for each phenotype from MWAS 1 and MWAS 2 were examined for overlap between early life environment and adult mental health. For the significant sites in MWAS 1, there were three annotated genes for preterm birth, having a young parent and population density, four annotated genes for low birth weight, 20 annotated genes for birth month, and 167 annotated genes for birth date. There were no annotated genes for the mental health phenotypes and therefore no overlap was observed with the early life environment phenotypes.

For significant sites in MWAS 2, there were three annotated genes for preterm birth, two annotated genes for low birth weight, and one annotated gene for having a young parent. As there were no annotated genes for the mental health phenotypes, there was no overlap of annotated genes with the early life environment phenotypes.

There were no enriched Gene Ontology or the KEGG gene sets for any of the phenotypes from either MWAS 1 or MWAS 2.

### Methylation profile scores

Set 1 results from MWAS 2 for the early life environment phenotypes were used to calculate profile scores for individuals in Set 2. The utility of these profile scores for predicting the same phenotype (Supplementary Table 4), MDD, or BRS (Table 3) in Set 2 was assessed. There was significant prediction of birth month using birth month profile scores (R^2^ = 0.002, *P* = 5.19 × 10-3) and of having a young parent using the profile scores from young parent (R^2^ = 0.013, *P* = 3.46 × 10-5). There was nominal prediction of MDD from preterm birth (R^2^ = 0.002, P = 0.034) and birth month (R^2^ = 0.002, *P* = 0.043), and nominal prediction of BRS from birth month (R^2^ = 0.003, P = 0.016). However, none of the profile scores predicting mental health phenotypes were significant after correction for multiple testing correction (*P* > 6.25 × 10^−3^).

### Blood cell type composition

In MWAS 1, there were 314 lead CpG sites (r^2^ > 0.1 within a 1 Mb window) associated with birth date (P < 7.01 × 10^−8^); however, in MWAS 2 there were no significant CpG sites for birth date. MWAS 2 included predicted blood cell type composition as a fixed effect, using haematological data obtained from the LBC. Access to the LBC was obtained and a MWAS run on five blood cell types. Of the 314 lead CpG sites from MWAS 1 in GS:SFHS, 220 were available in the LBC. The QQ-plots for the observed and expected P-values and λ for these 220 sites in LBC are provided in Supplementary Figures 17-21 for each blood cell type. Examination of these plots revealed no inflation of P-values for basophil, eosinophil, and monocyte counts. However, there was noticeable inflation for neutrophil (50 out of 220 sites with P < 0.05; λ = 2.503) and lymphocyte (98 out of 220 sites with P < 0.05; λ = 5.841) counts. Similarly, creating methylation profile scores using the MWAS 1 effect sizes for birth date in GS:SFHS (Supplementary Table 5), revealed significant prediction of neutrophil (*P* = 7.81 × 10^−4^) and lymphocyte counts (*P* = 3.45 × 10^−7^) in the LBC. These profile scores captured 0.8% and 1.9% of the phenotypic variance of the neutrophil and lymphocyte counts, respectively.

## Discussion

The impact of early life environments on later life is of critical importance across multiple clinical and research domains. We sought to examine eight early life environments experienced around the time of birth and quantify: (1) their impact on mental health in adulthood; (2) whether they led to detectable changes to the methylome in to adulthood; and finally (3) whether there was any shared association on the methylome between early life environments and adult mental health.

Previous studies have reported an effect of extreme preterm (gestation length less than 28 weeks) on mental health [8, 54]. The population-based GS:SFHS cohort had four participants (out of 3,134) born at less than 28 weeks gestation length and therefore a 37-week threshold for a preterm birth was used. Three CpG sites were associated with preterm birth in both the MWAS analyses: cg00725333, cg17668848, and cg24329141. These three sites mapped to PRICKLE2 (Prickle Planar Cell Polarity Protein 2), *HHLA2* (Human Endogenous Retrovirus-H Long Terminal Repeat-Associating Protein 2), and *ABI1* (Abl Interactor 1) protein coding genes, respectively. Homologs of PRICKLE2 have been associated with embryonic development in mice [56, 57] and drosophila [58]. In humans, PRICKLE2 has been associated with myelomeningocele [59], a severe form of spina bifida. Myelomeningocele compromises the development of the spine and spinal cord in the womb and has been associated with preterm birth [60]. *HHLA2* regulates T-cell function [61] with increased expression observed in osteosarcoma tumours [62], however its role in gestation length is unclear. Knockouts of ABI1 in mice have demonstrated it to be essential for embryonic development, survival [63] and placental development [64]. *PRICKLE2* and *ABI1* have plausible roles in gestation length with altered methylation of proximal CpG sites into adulthood. A methylome-wide association study conducted using preterm infants [65] revealed a different set of associated CpG sites to those observed here in adulthood.

Similar to preterm birth, published studies of birth weight and mental health report that more extreme birth weight thresholds identify stronger associations with poorer mental health [66, 67]. In the current study, a sex and gestational-age adjusted measure of birth weight was used increasing the distinction with the preterm birth phenotype. Five CpG sites were associated with birth weight in MWAS 1 of which two were also significant in MWAS 2. The two sites that were significant in both MWAS annotated to genes important for the development of the sensory systems: SOBP (Sine Oculis Binding Protein Homolog) and *RPGRIP1* (Retinitis Pigmentosa GTPase Regulator Interacting Protein). SOBP has been implicated in the embryonic development of the mouse cochlea [68]. In a meta-analysis of DNA methylation data in neonates for low birth weight, a CpG site annotated to the SOBP gene was also significant after correcting for false discovery rate (but not after Bonferroni correction) [69]. The critical role of *RPGRIP1* in the remodelling of rod photoreceptors has been demonstrated in humans [70]. The impact of low birth weight on the long-term expression of CpG sites requires further investigation, certainly as low birth weight has been associated with both hearing impairment [71] and ophthalmic deficits [72]. cg21803443 on chromosome 7 was significant in MWAS 1 and annotated to the GRM8 (Glutamate Metabotropic Receptor 8) protein-coding gene. Küpers et al. [69] also identified a CpG site (cg15908975, P = 4.52 × 10^−7^ in a European meta-analysis) close to *GRM8* associated with birth weight. GRM8 is involved in the inhibition of the cyclic AMP cascade influencing glutamatergic neurotransmission and had an association with depression (*P* = 1.80 × 10^−12^) in a genome-wide association meta-analysis [73]. However, there was no predictive ability of a methylation profile score for low birth weight to predict either MDD or BRS. A previous analysis of birth weight as a continuous trait in GS:SFHS identified one significant CpG site (cg00966482) [74], however this site was not significant in either MWAS 1 (P = 0.22) or MWAS 2 (P = 0.53).

Schnittker [12] suggests that the role of seasonality of birth on mental health was more prevalent in the early part of the 20^th^ century and was partly attributable to poorer prenatal nutrition across the winter months. However, Disanto et al. [10] analysed post-1950 data from England and reported an effect of time of year of birth on schizophrenia, bipolar disorder and to a lesser extent recurrent MDD. In the current study, there was nominal evidence for an effect of time of year of birth on BRS and nominal prediction of both MDD and BRS using a profile score for birth month calculated from DNA methylation data. The greatest difference between the MWAS 1 and MWAS 2 results was for the time of year of birth phenotypes. Additional prediction into the LBC, using methylation profile scores, demonstrated that this divergence is likely due to differing blood cell type composition (for neutrophils and lymphocytes) between those born in the winter and those born in the summer. Altered gene expression due to seasonality has been reported previously [75], with the time of year of birth also influencing neonatal immune development [76] and thymic output [77]. The current work provides further substantial evidence that time of year of birth impacts blood cell type composition. This impact is detectable throughout the life course and needs to be accounted for in future research on traits influenced by seasonality of birth.

In the present study, there was no effect of having a young parent on mental health. However, a much larger study (2.9M individuals) on Danish participants found an increased risk of mood disorders (International Classification of Disease codes F30 – 39) for those with teenage mothers (incidence risk ratio = 1.35 [95% CI = 1.30 – 1.40]) and those with teenage fathers (incidence risk ratio = 1.20 [95% CI = 1.13 – 1.27]) [13]. The *MTNR1B* (Melatonin Receptor 1B) protein coding gene was annotated to a CpG site that was associated with having a young parent in both MWAS 1 and MWAS 2 and the *PARD6G-AS1* (PARD6G Antisense RNA 1) RNA gene was annotated to an associated CpG site in MWAS 1. *MTNR1B* has been associated with type 2 diabetes [78, 79] and differential expression in *PARD6G-AS1* has been implicated in transient neonatal diabetes [80]. This suggests a putative link between having a young parent and diabetes, but further work is required to validate any association. The methylation profile for having a young parent from Set 1 had significant prediction of having a young parent in Set 2, suggesting replication across Sets.

The lone parent phenotype in the current study did not account for whether the mother or father was absent, custody arrangements, separation during infancy, or the degree of communication with the absent parent. A large study of 184,496 school-aged children found that life satisfaction was decreased when either parent was absent, but where the father was absent the effect was attenuated when accounting for perceived family affluence [81]. A follow-up to this study, incorporating additional cohorts, also reported similar effects [82]. The present study found no effect of having a lone parent on MDD or BRS and there were no significant associations with any of the CpG sites analysed.

Two correlated early life phenotypes, urban environment and population density, were studied to measure the effect of geographical environment at birth. There was no effect of either early life phenotype on MDD or BRS and there were no associated CpG sites. Urbanicity has been associated with depression [83, 84], although no association has been observed in low- and middle-income countries [85] and the United States [86]. Population density has also been shown to have limited impact on depression in Turin, Italy [87]. The contradictory findings and the lack of observable effects here may be due to the multiple factors incorporated in these phenotypes, such as pollution [88] and socio-economic status [89], but may be offset by access to mental health services.

Published DNA methylation analysis of MDD have typically been conducted on relatively small samples. The two CpG sites associated with MDD in the current study did not annotate to known protein coding genes. These two sites were outside of the ten differentially methylated regions for major depression identified by Roberson-Nay et al. [90] (39 major depression cases and 111 controls) and located away from the locations identified for MDD by Oh et al. [91] (103 cases and 97 controls) and Starnawska et al. [92] (724 individuals assessed for depression symptomatology score). A longitudinal methylome-wide association study of MDD (199 cases and 382 in remittance) identified six plausible CpG sites based on function [93], although none of the sites remained significant after applying correction for multiple testing. A further study of profile scores for MDD in GS:SFHS found that prediction of MDD in an independent subset was possible, but was reliant on capturing lifestyle factors associated with MDD [38]. An association meta-analysis (7,948 individuals) identified 20 CpG sites with a suggestive association (*p* < 1 × 10^−5^) with depressive symptoms [23], of which one was nominally significant (*p* = 0.048) in a replication cohort of 3,308 individuals. None of these 20 suggestive sites were close to either of the associated sites observed in the current study. To reach replicable findings for CpG sites associated with MDD it is likely that larger sample sizes will be needed as has been demonstrated in genome-wide associations studies [73].

The initial methodology of the current study was to use a single linear regression, MWAS 1. It was only after observing genomic inflation in the signal for the time of year of birth phenotypes that MWAS 2 was also included. MWAS 2 accounts for methylome-wide correlational structure and makes additional adjustment for predicted cell type composition. The non-significance of any of the associated MWAS 1 variants in MWAS 2, suggests cell type composition as the likely cause of the inflation, which was confirmed by our analysis in the LBC. Therefore, correcting M-values for cell type composition using the Houseman algorithm [43] may not be adequate, as there remained a detectable effect of cell type composition in the GS:SFHS data. Furthermore, if we had only run MWAS 2 we would not have observed the effect of time of year on birth on cell type composition. In future, it may be preferable to run multiple regression models on DNA methylation data.

The results reported here are based on a single European population and their applicability to other countries and ancestries is unknown. GS:SFHS is a family-based sample drawn from the general population. Therefore, the more extreme phenotypes that have been analysed in the published literature would not have provided adequate power in the studied cohort. There was also no measure of childhood abuse or neglect collected in GS:SFHS and those phenotypes may provide additional avenues for investigation using a similar methodology to that used here. Finally, the DNA methylation data analysed was obtained from blood and the analysis of other tissue samples may reveal additional associations with the phenotypes examined.

In conclusion, there were plausible CpG sites associated with preterm birth, low birth weight and having a young parent in both MWAS 1 and MWAS 2. Further, one of the more interesting findings was the long-term impact of the time of year of birth on the blood cell type composition for neutrophils and lymphocytes. It was not possible to predict either MDD or BRS from methylation profile scores calculated from early life phenotypes. Although, there was significant predictive ability across the two sets of GS:SFHS data for birth month and having a young parent suggesting the reliability of the results for those phenotypes.

## Data Availability

Methylome-wide summary statistics calculated from MWAS 1 and MWAS 2 for each of the assessed phenotypes will be made publicly available following publication of the peer-reviewed manuscript

## Code and Data availability

Methylome-wide summary statistics calculated from MWAS 1 and MWAS 2 for each of the assessed phenotypes are available from: TBC

## Author contributions

D.M.H., A.M.McI., M.M., and C.M.L. conceived the research project. D.M.H, I.J.D, D.P, A.C, L.A, and A.M.McI determined the early life phenotypes used. M.C.B, C.A, R.M.W, and K.L.E applied quality control to the Generation Scotland DNA methylation data. D.M.H ran the analyses. O.P, R.A, M.C.B, B.J., M.J.A., P.F.S., A.M.McI., N.R.W. and C.M.L. provided expertise of methylome-wide association study methodology and statistical analysis. All authors commented on the manuscript.

## Conflicts of interest

A.M.McI. has received research support from Eli Lilly and Company, Janssen and the Sackler Trust and speaker fees from Illumina and Janssen. C.M.L is a member of the Myriad Neuroscience R&D SAB.

## Acknowledgements

Generation Scotland received core support from the Chief Scientist Office of the Scottish Government Health Directorates [CZD/16/6] and the Scottish Funding Council [HR03006] and is currently supported by the Wellcome Trust [216767/Z/19/Z]. Genotyping of the GS:SFHS samples was carried out by the Genetics Core Laboratory at the Edinburgh Clinical Research Facility, University of Edinburgh, Scotland and was funded by the Medical Research Council UK and the Wellcome Trust (Wellcome Trust Strategic Award “STratifying Resilience and Depression Longitudinally” (STRADL) Reference 104036/Z/14/Z). We are grateful to all the families who took part, the general practitioners and the Scottish School of Primary Care for their help in recruiting them, and the whole Generation Scotland team, including interviewers, computer and laboratory technicians, clerical workers, research scientists, volunteers, managers, receptionists, healthcare assistants and nurses. Ethics approval for the study was given by the NHS Tayside committee on research ethics (reference 05/S1401/8).

LBC1921 funding has been received from the UK’s Biotechnology and Biological Sciences Research Council (BBSRC) (15/SAG09977, wave 1), a Royal Society-Wolfson Research Merit Award to IJD (wave 2), the Chief Scientist Office (CSO) of the Scottish Government’s Health Directorates (CZG/3/2/79, post-wave 1 questionnaire study; CZB/4/505, wave 3; ETM/55, wave 4), and the UK’s Medical Research Council (MRC) Centenary Early Career Award to Dr Tom Booth (wave 5). Funding for LBC1936 has been received from Research Into Ageing (Programme grant 251; wave 1), and Age UK (Disconnected Mind Programme grant) and the UK’s Medical Research Council (G0701120, wave 2; G1001245, wave 3; MR/M013111/1, wave 4). The Alzheimer Scotland Dementia Research Centre funded LBC1936 dementia ascertainment. BBSRC funded whole-genome sequencing of both cohorts. Wellcome, the University of Edinburgh, the University of Queensland, and Age UK funded DNA methylation analysis in both cohorts.

D.M.H is supported by a Sir Henry Wellcome Postdoctoral Fellowship (Reference 213674/Z/18/Z) and a 2018 NARSAD Young Investigator Grant from the Brain & Behavior Research Foundation (Ref: 27404). A.M.McI is supported by Wellcome Trust (104036/Z/14/Z and 216767/Z/19/Z) and UKRI MRC funding (MC PC 17209 and MR/S035818/1). C.A is supported by the Medical Research Council (MRC) UK (grants MC_PC_U127592696 and MC_PC_U127561128). I.J.D. was supported by the Centre for Cognitive Ageing and Cognitive Epidemiology, which was funded by the Medical Research Council and the Biotechnology and Biological Sciences Research Council (MR/K026992/1). R.E.M is supported by Alzheimer’s Research UK major project grant ARUK-PG2017B-10. N.R.W. acknowledges NMHRC grants 1078901 and 1087889. L.A is the Mental Health Leadership Fellow for the ESRC. C.M.L acknowledges MRC grant MR/N015746/1. This investigation represents independent research part-funded by the National Institute for Health Research (NIHR) Biomedical Research Centre at South London and Maudsley NHS Foundation Trust and King’s College London. The views expressed are those of the authors and not necessarily those of the NHS, the NIHR or the Department of Health and Social Care.

## Notes

### Author Declarations

Ethics approval for Generation Scotland was given by the NHS Tayside committee on research ethics (reference 05/S1401/8).

