## Supplementary Figures for "Methylome-wide association study of early life stressors and adult mental health reveals a relationship between birth date and cell type composition in blood"

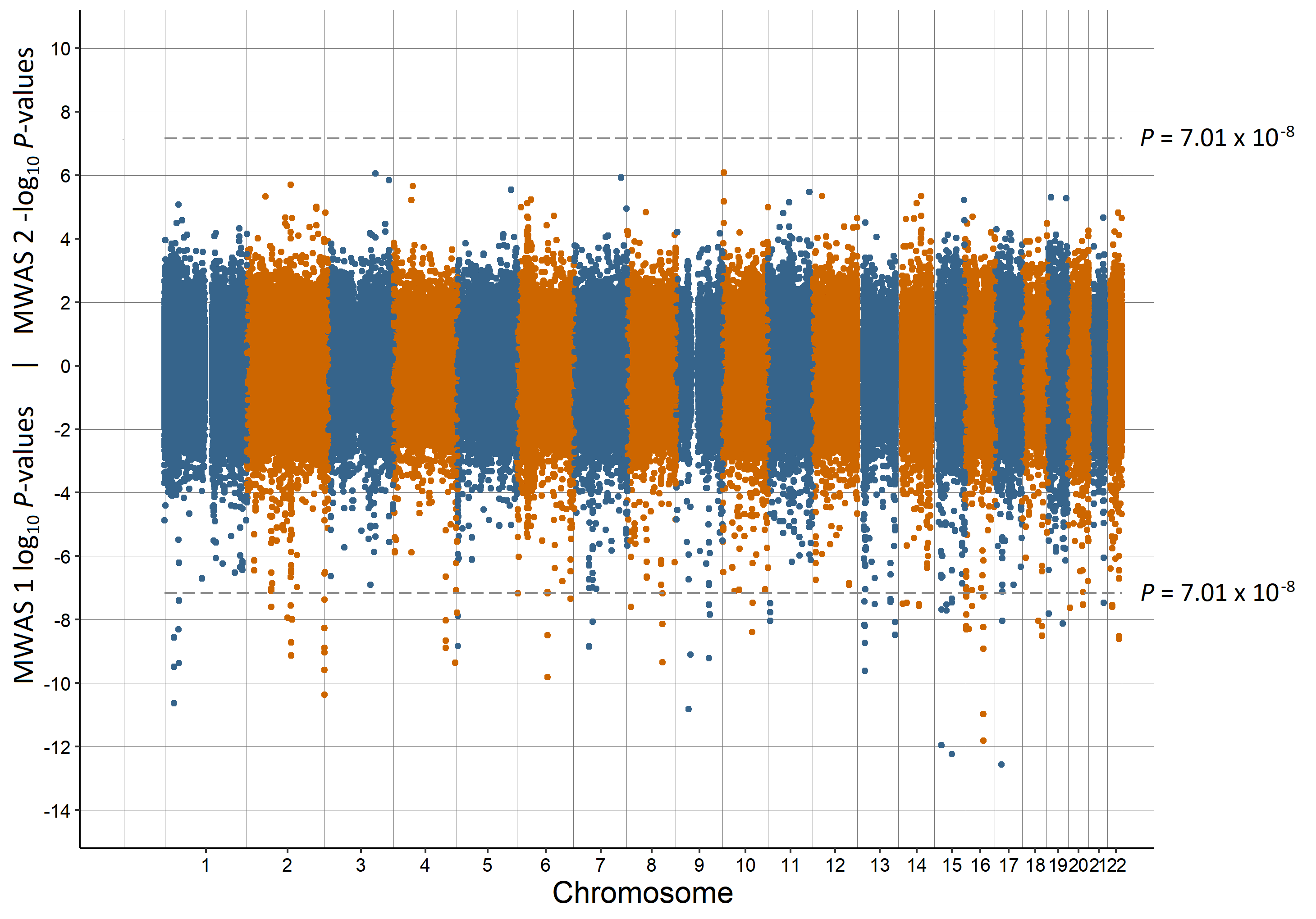

Supplementary Figure 1. Miami plot of the observed *P*-values of each CpG site for an association with birth month

Log_10_ *P*-values are shown for MWAS 1 and -log_10_ *P*-values are shown for MWAS 2. The dotted lines indicate methylome-wide significance (*P* = 7.01 × 10^-8^)

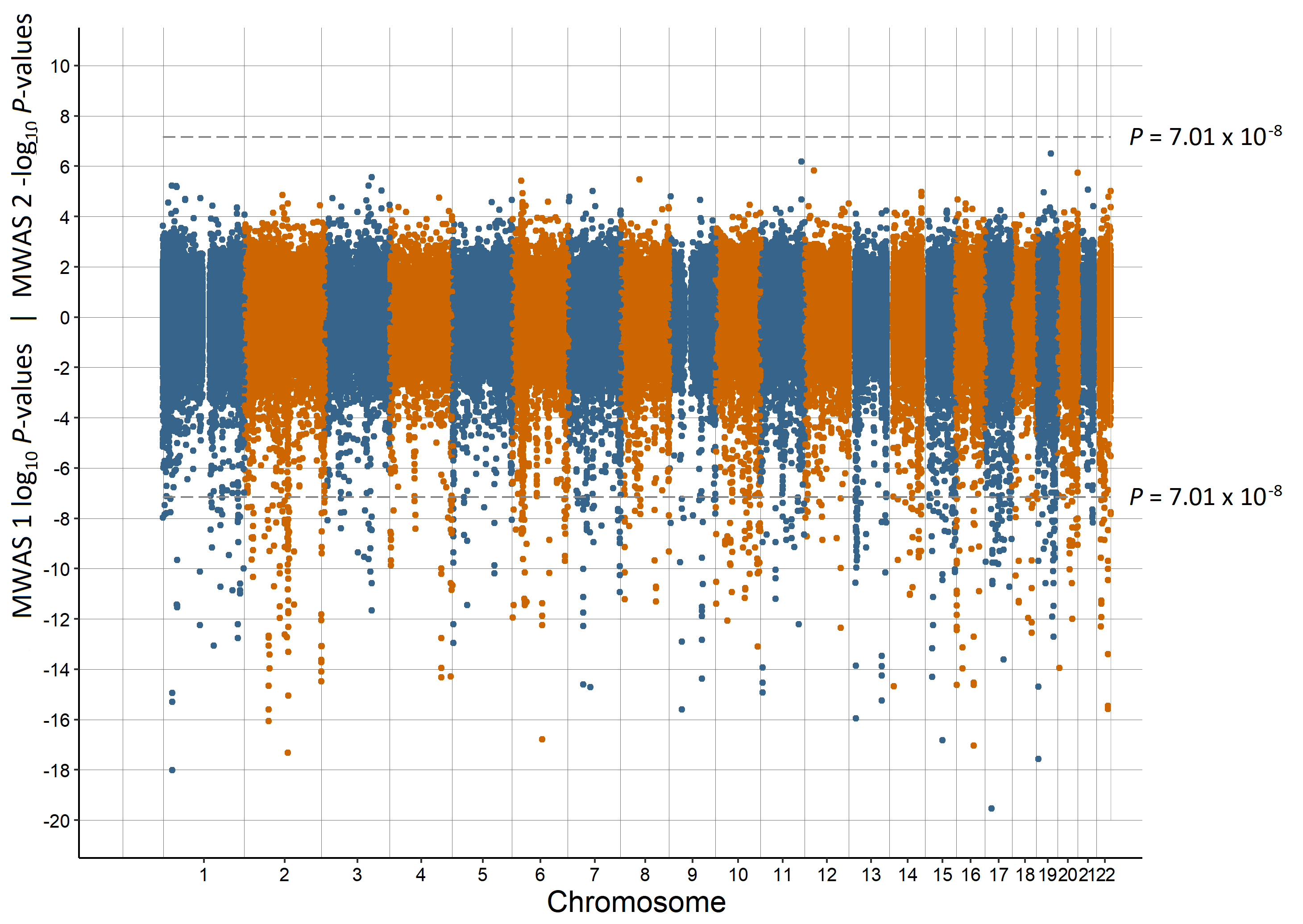

Supplementary Figure 2. Miami plot of the observed *P*-values of each CpG site for an association with birth date

Log_10_ *P*-values are shown for MWAS 1 and -log_10_ *P*-values are shown for MWAS 2. The dotted lines indicate methylome-wide significance (*P* = 7.01 × 10^-8^)

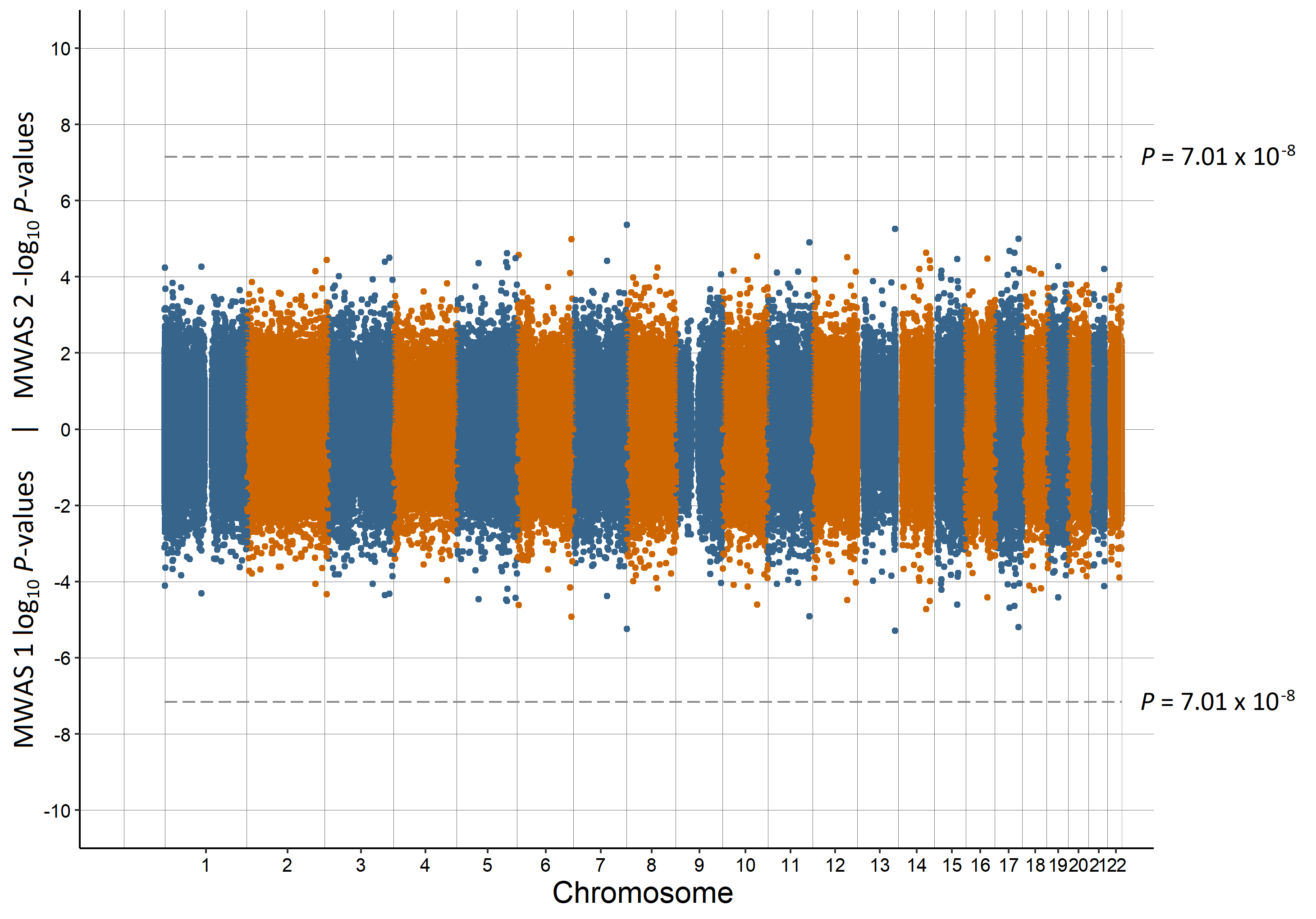

Supplementary Figure 3. Miami plot of the observed *P*-values of each CpG site for an association with having a lone parent

Log_10_ *P*-values are shown for MWAS 1 and -log_10_ *P*-values are shown for MWAS 2. The dotted lines indicate methylome-wide significance (*P* = 7.01 × 10^-8^)

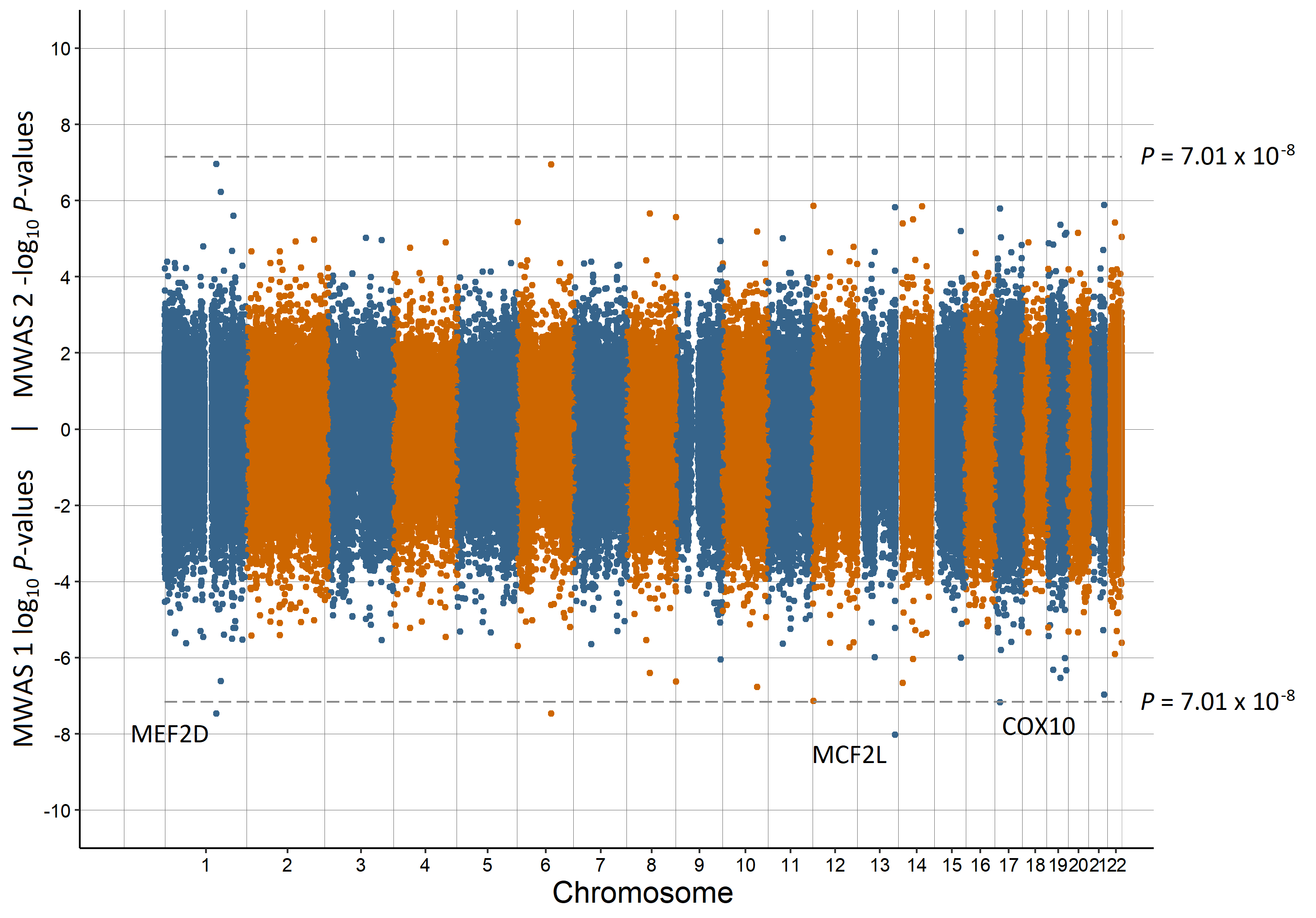

Supplementary Figure 4. Miami plot of the observed *P*-values of each CpG site for an association with population density

Log_10_ *P*-values are shown for MWAS 1 and -log_10_ *P*-values are shown for MWAS 2. The dotted lines indicate methylome-wide significance (*P* = 7.01 × 10^-8^). The annotation of genes for significant sites is reported by missMethyl for MWAS 1.

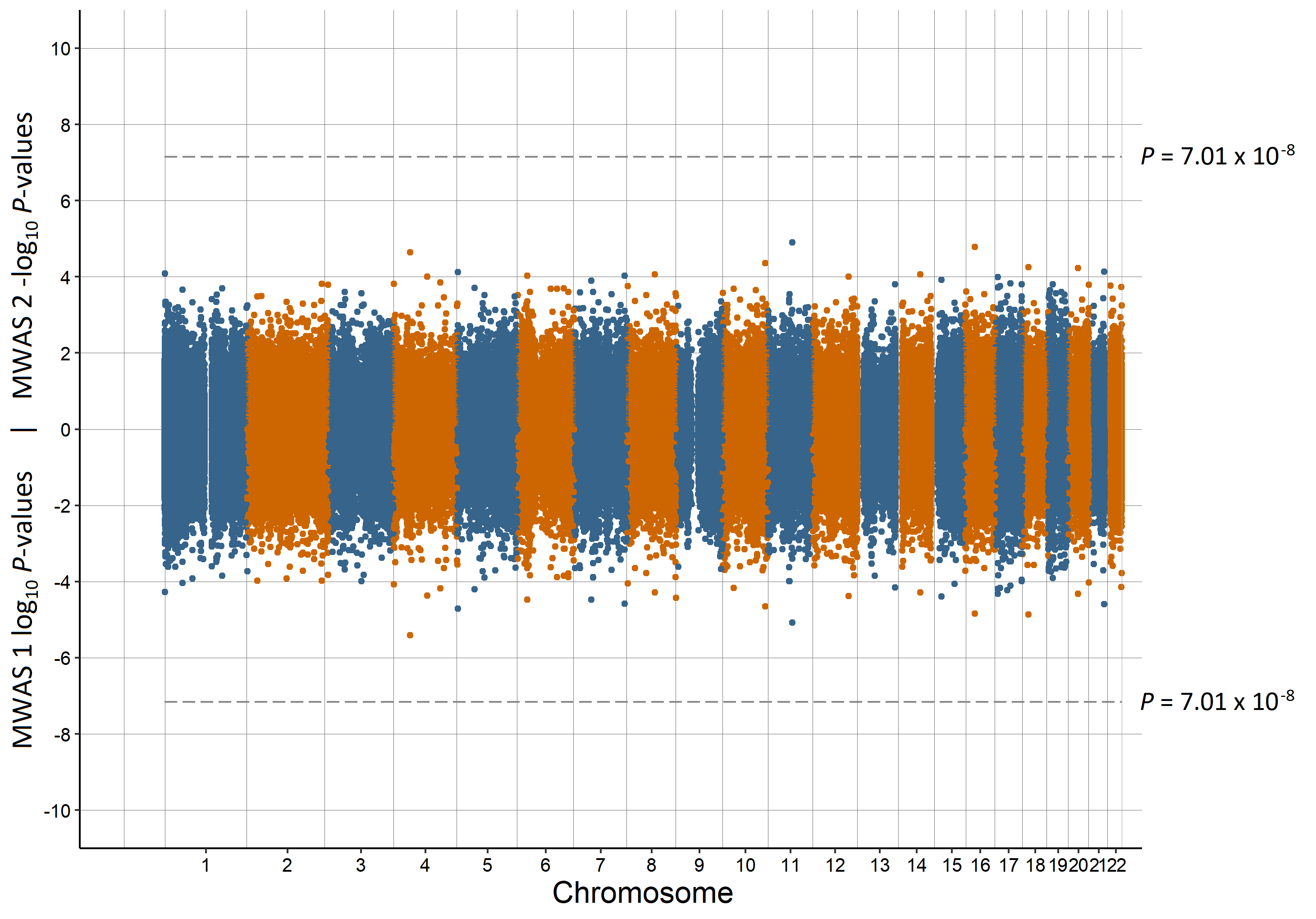

Supplementary Figure 5. Miami plot of the observed *P*-values of each CpG site for an association with urbanicity

Log_10_ *P*-values are shown for MWAS 1 and -log_10_ *P*-values are shown for MWAS 2. The dotted lines indicate methylome-wide significance (*P* = 7.01 × 10^-8^)

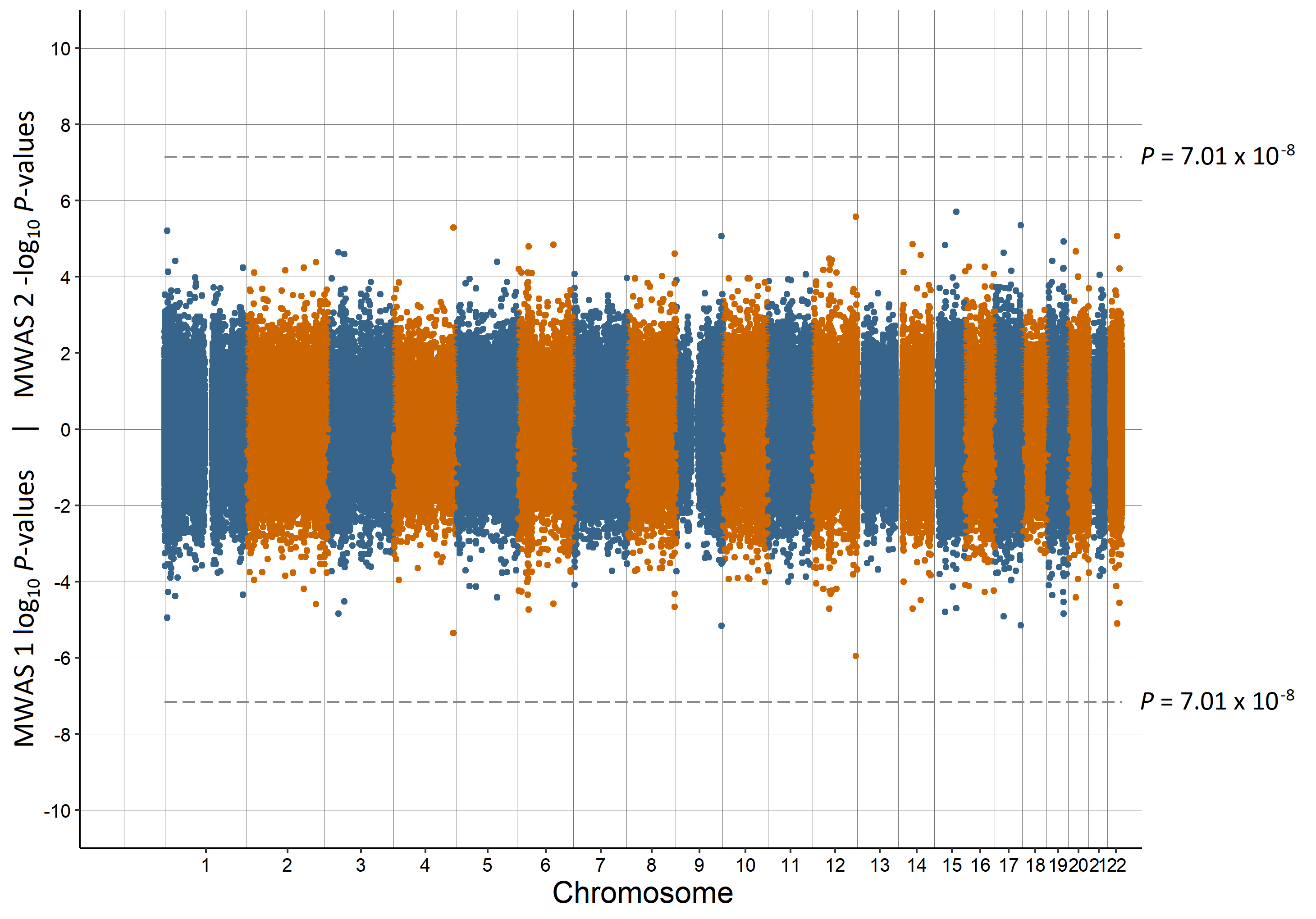

Supplementary Figure 6. Miami plot of the observed *P*-values of each CpG site for an association with brief resilience scale

Log_10_ *P*-values are shown for MWAS 1 and -log_10_ *P*-values are shown for MWAS 2. The dotted lines indicate methylome-wide significance (*P* = 7.01 × 10^-8^)

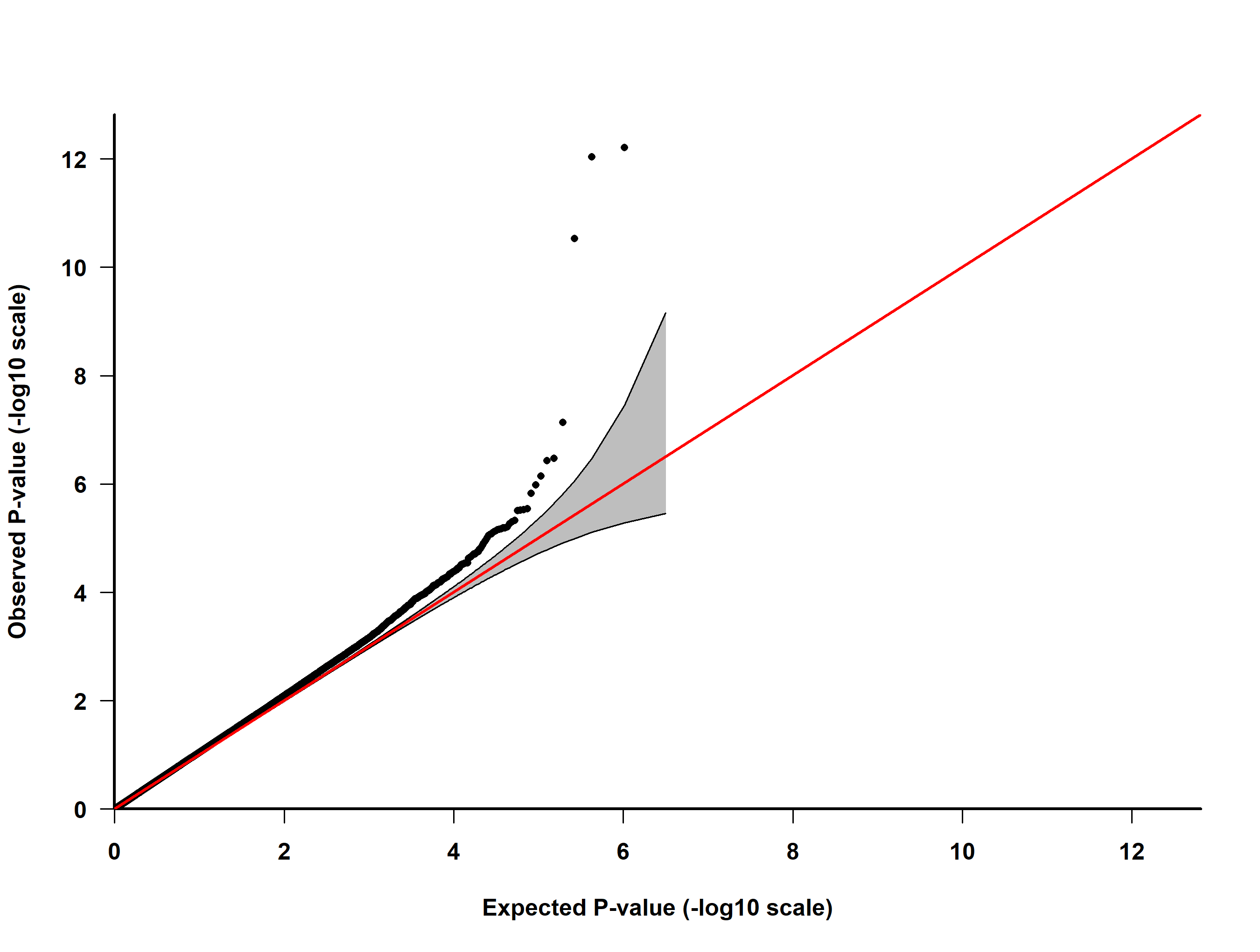

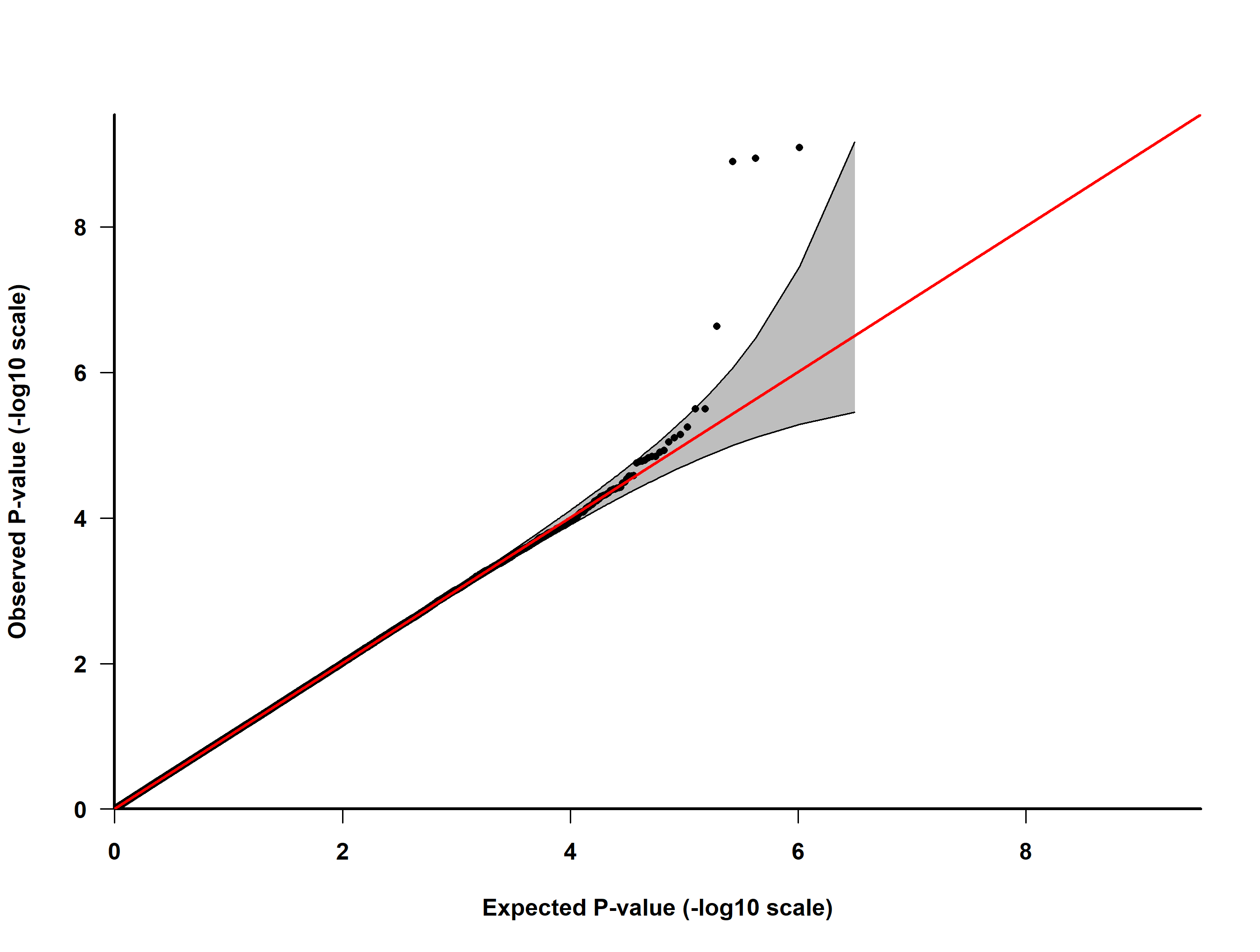

B

A

Supplementary Figure 7. QQ-plot of observed and expected *P*-values for preterm birth in MWAS 1 (A) and MWAS 2 (B)

The straight line is where the observed *P*-values match those expected and the shaded area is the 95% confidence interval. Genomic inflation: MWAS 1 = 1.070, MWAS 2 = 1.018

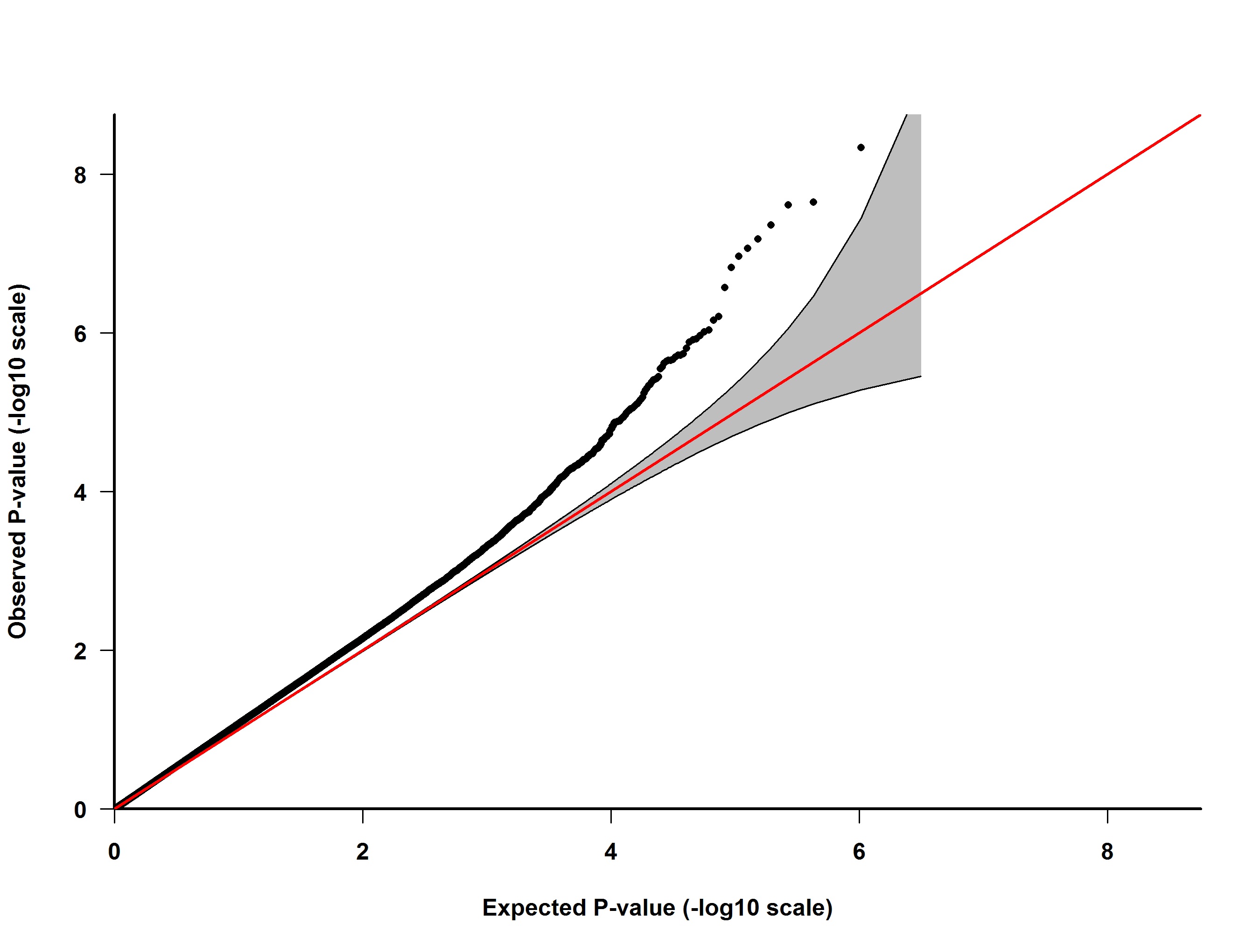

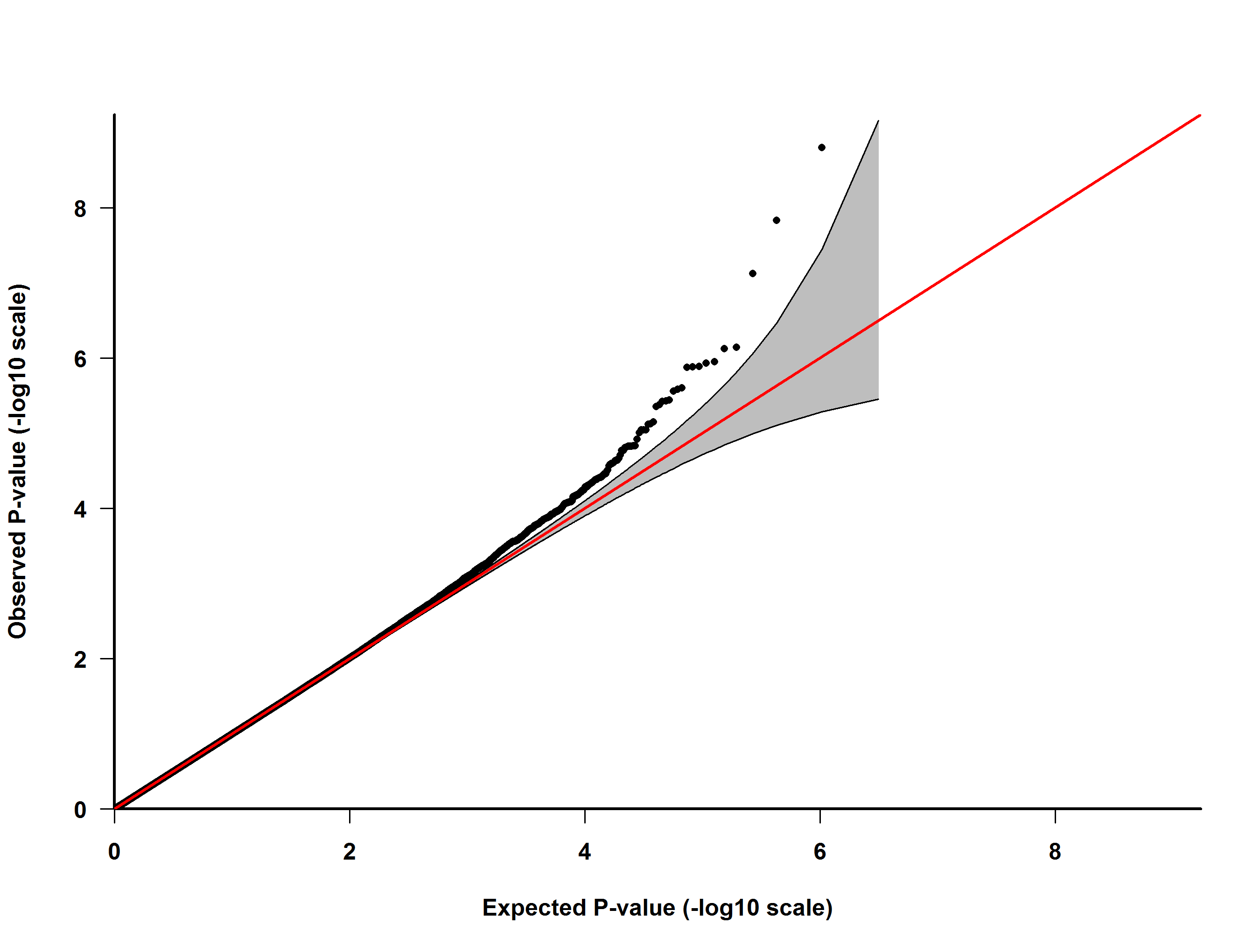

B

A

Supplementary Figure 8. QQ-plot of observed and expected *P*-values for low birth weight in MWAS 1 (A) and MWAS 2 (B)

The straight line is where the observed *P*-values match those expected and the shaded area is the 95% confidence interval. Genomic inflation: MWAS 1 = 1.117, MWAS 2 = 0.999

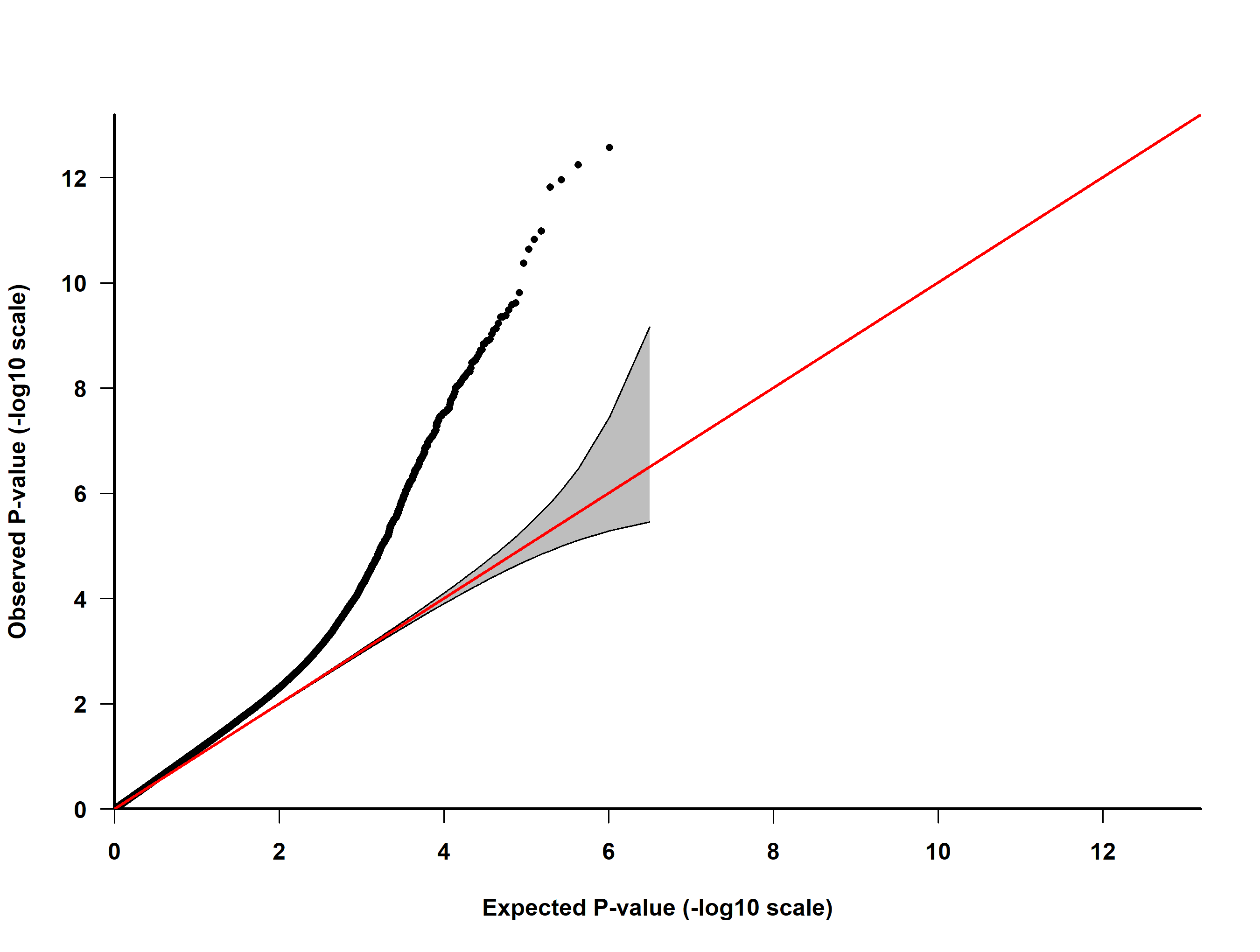

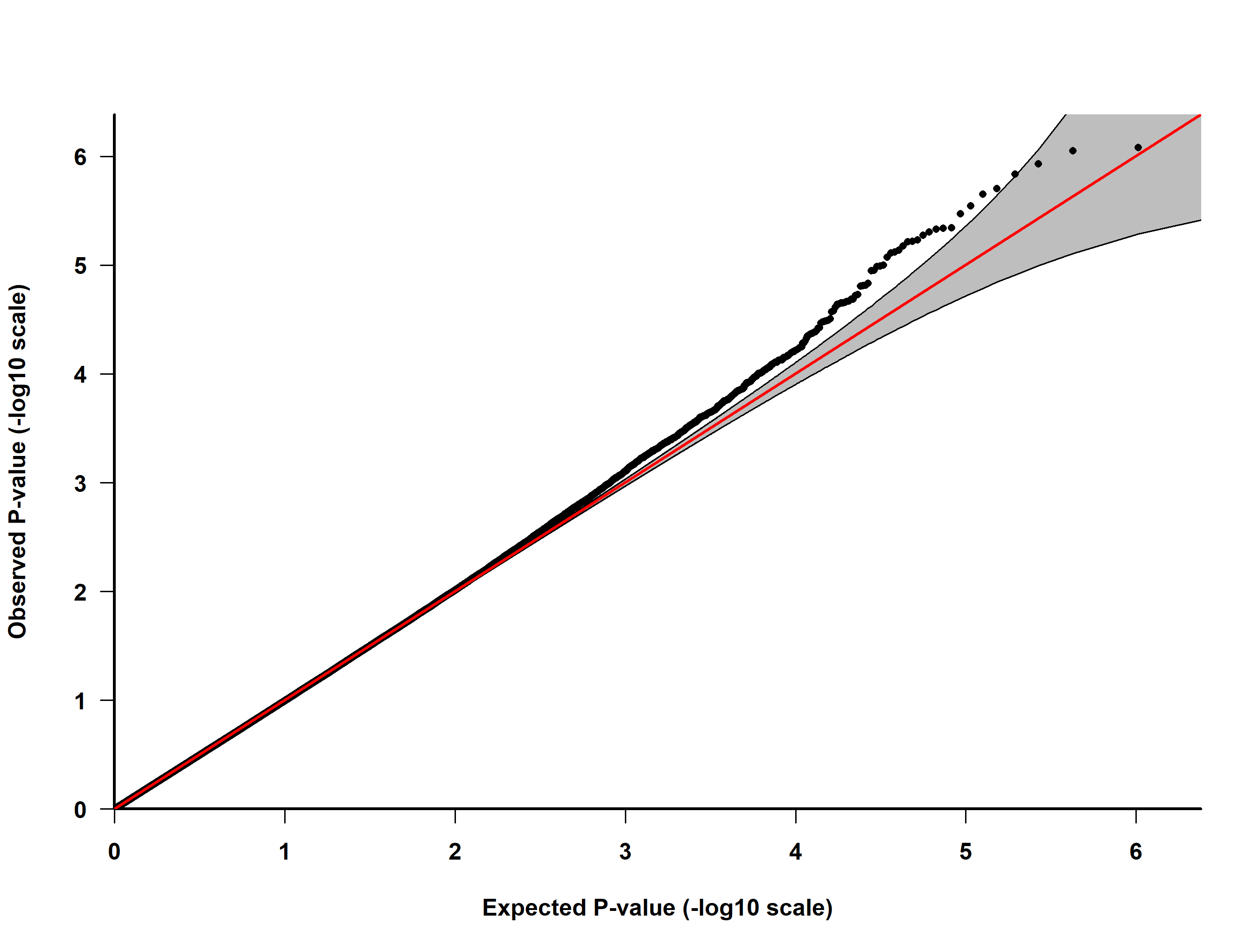

B

A

Supplementary Figure 9. QQ-plot of observed and expected *P*-values for birth month in MWAS 1 (A) and MWAS 2 (B)

The straight line is where the observed *P*-values match those expected and the shaded area is the 95% confidence interval. Genomic inflation: MWAS 1 = 1.166, MWAS 2 = 0.989

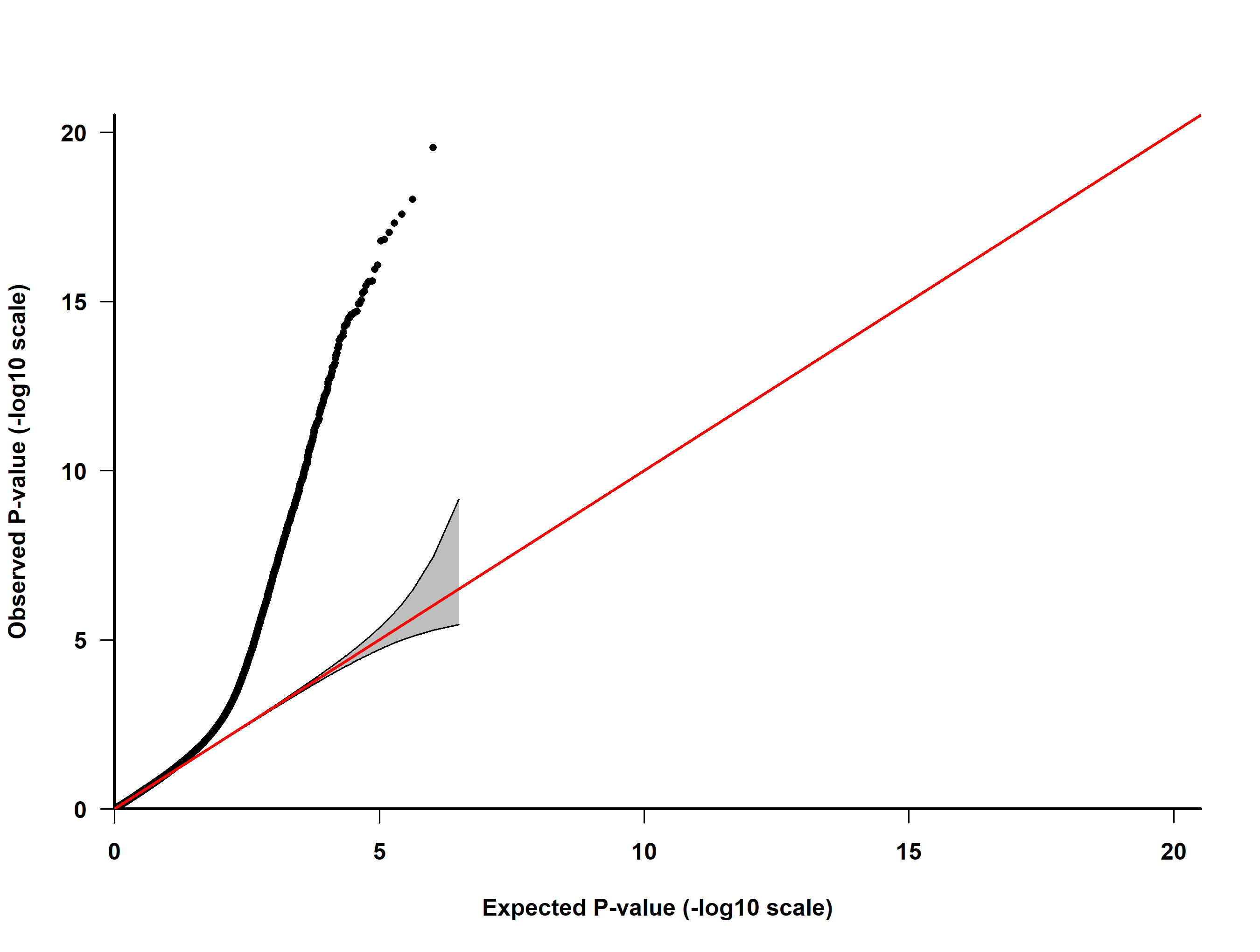

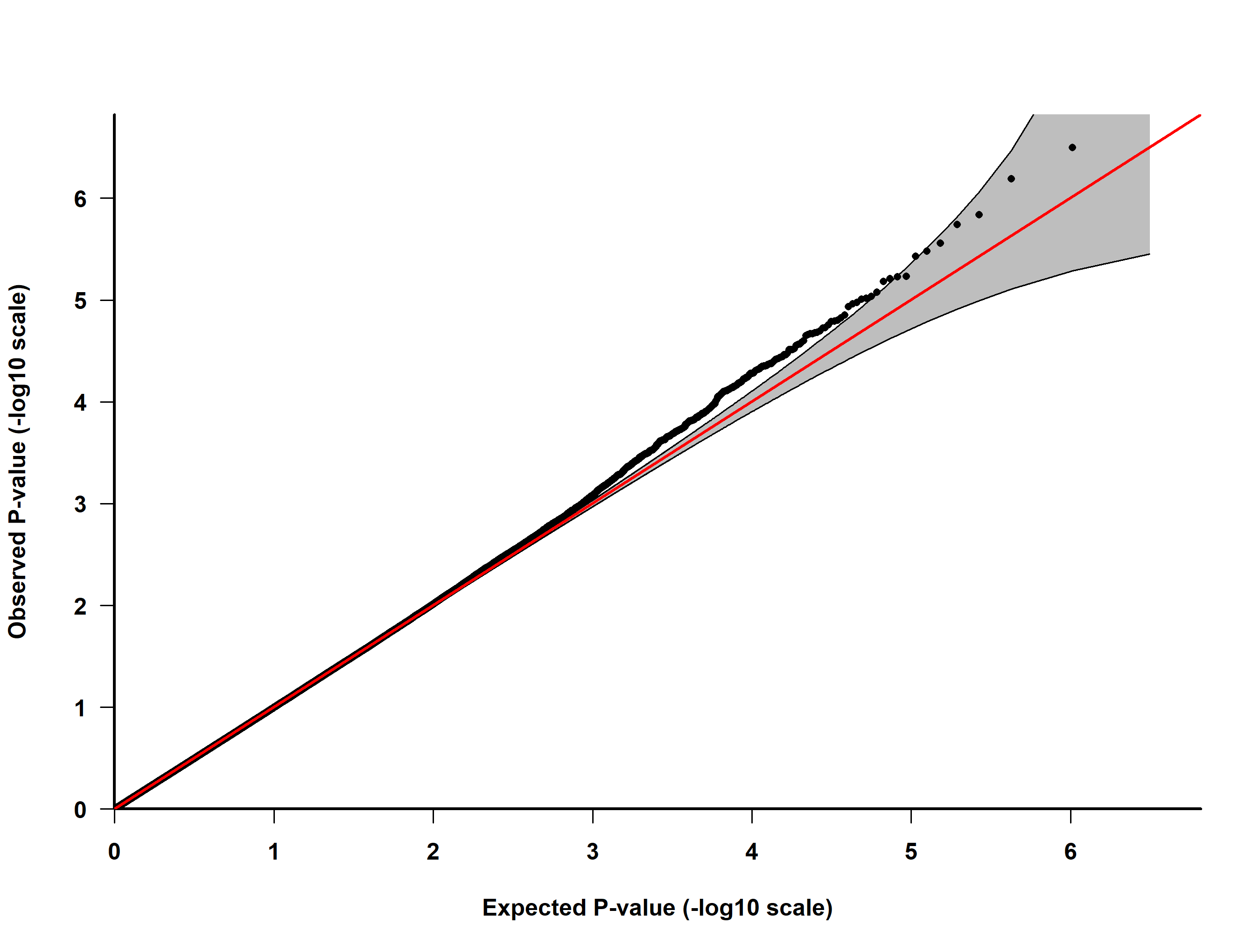

B

A

Supplementary Figure 10. QQ-plot of observed and expected *P*-values for birth date in MWAS 1 (A) and MWAS 2 (B)

The straight line is where the observed *P*-values match those expected and the shaded area is the 95% confidence interval. Genomic inflation: MWAS 1 = 1.010, MWAS 2 = 0.989

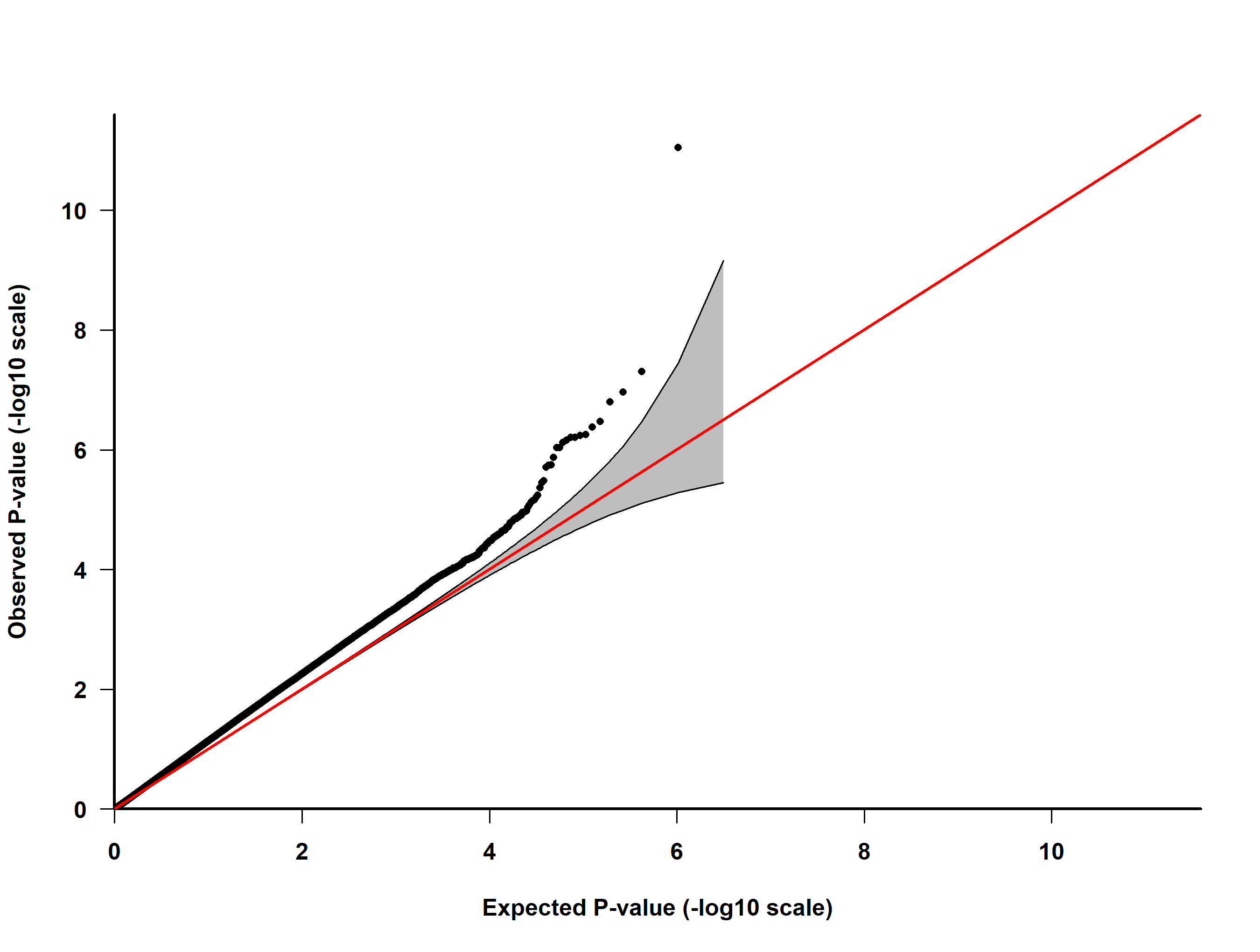

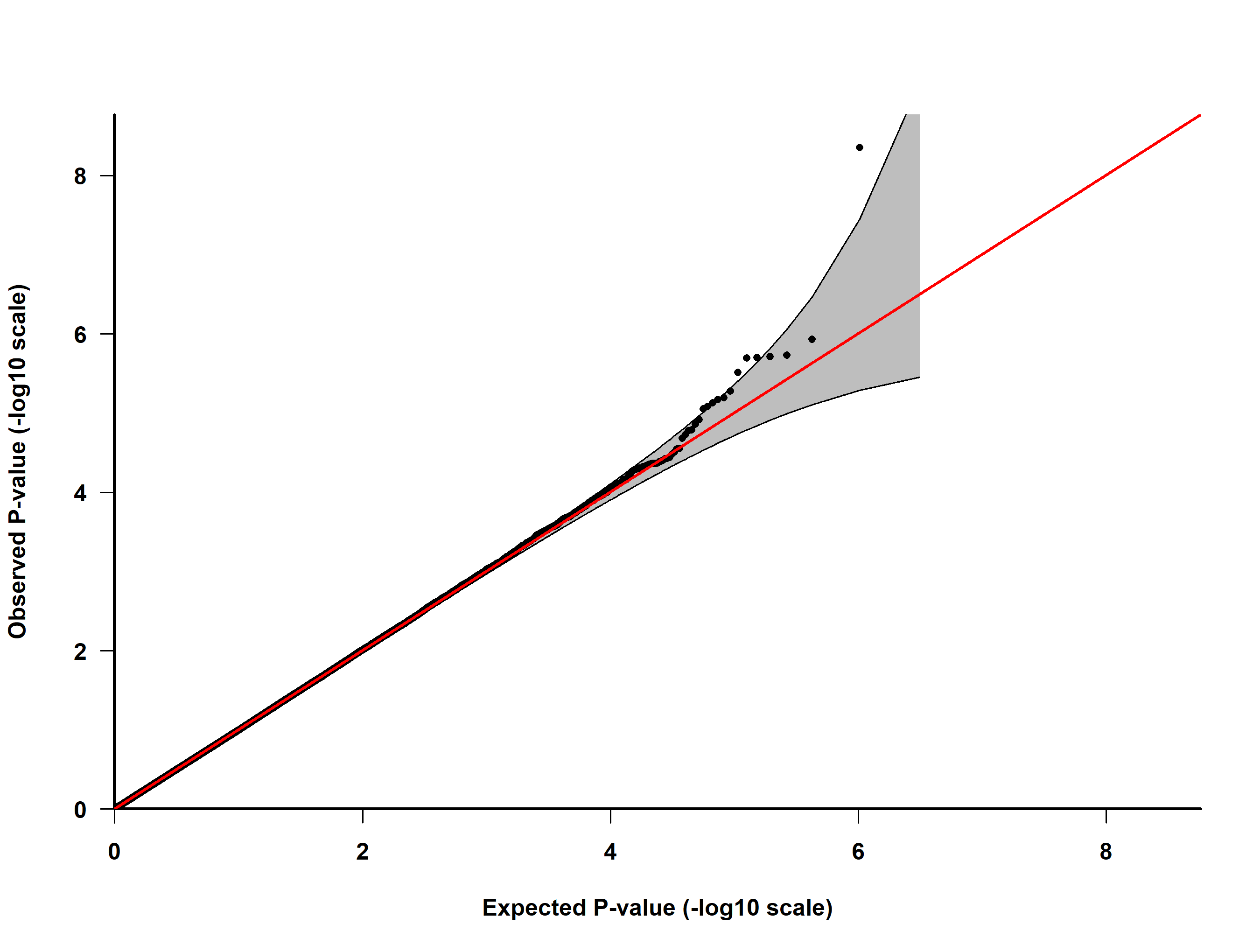

B

A

Supplementary Figure 11. QQ-plot of observed and expected *P*-values for having a young parent in MWAS 1 (A) and MWAS 2 (B)

The straight line is where the observed *P*-values match those expected and the shaded area is the 95% confidence interval. Genomic inflation: MWAS 1 = 1.207, MWAS 2 = 0.997

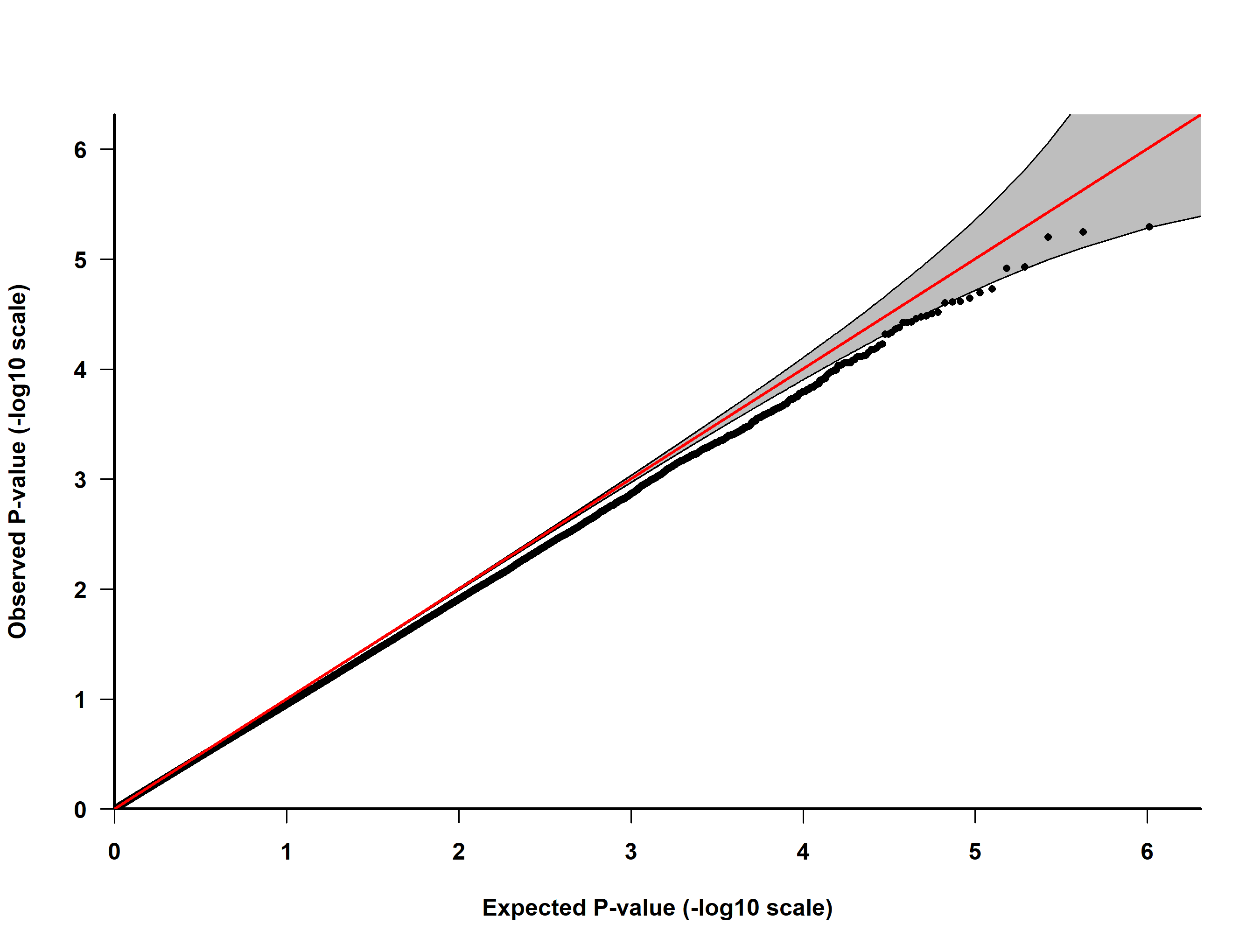

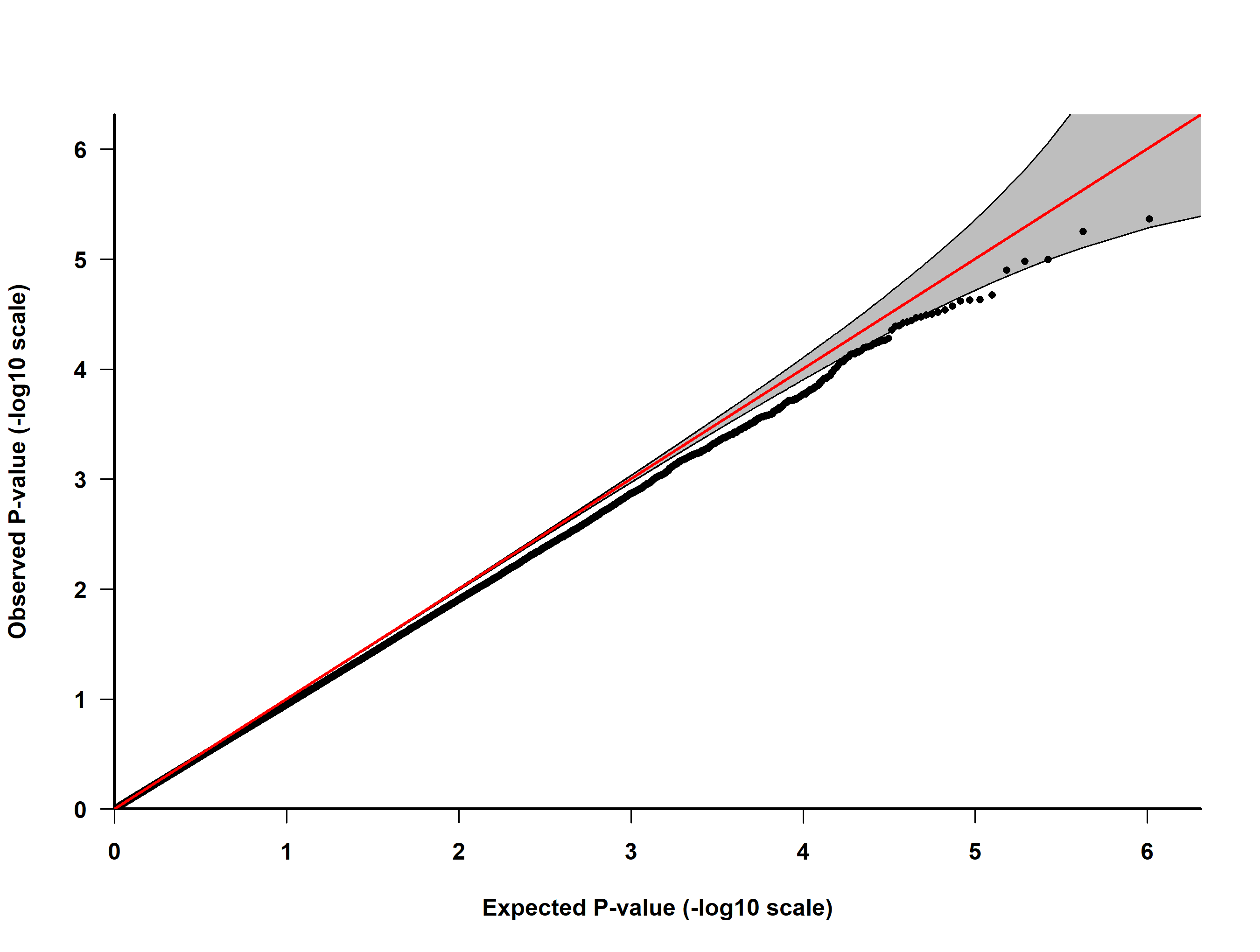

B

A

Supplementary Figure 12. QQ-plot of observed and expected *P*-values for having a lone parent in MWAS 1 (A) and MWAS 2 (B)

The straight line is where the observed *P*-values match those expected and the shaded area is the 95% confidence interval. Genomic inflation: MWAS 1 = 0.932, MWAS 2 = 0.932

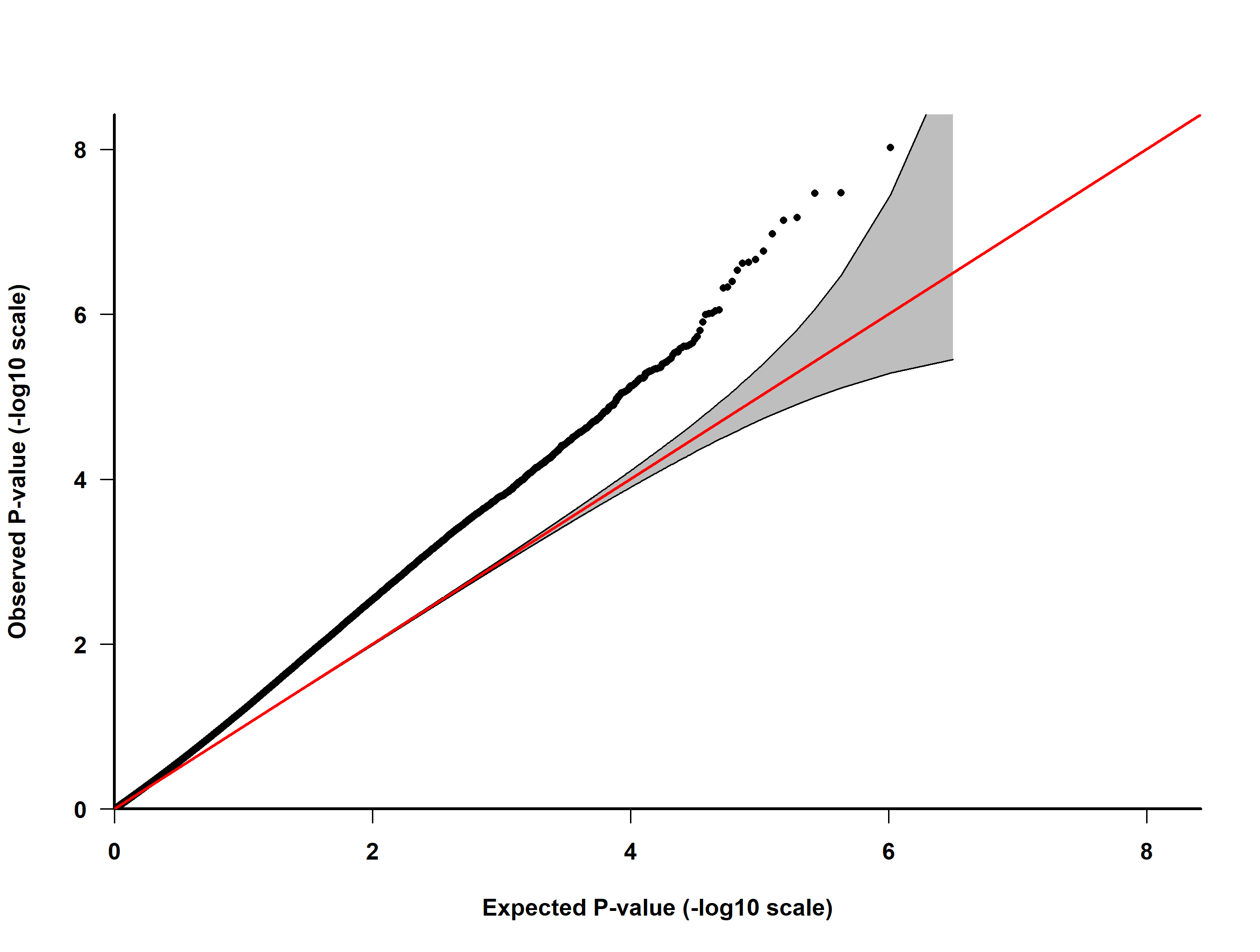

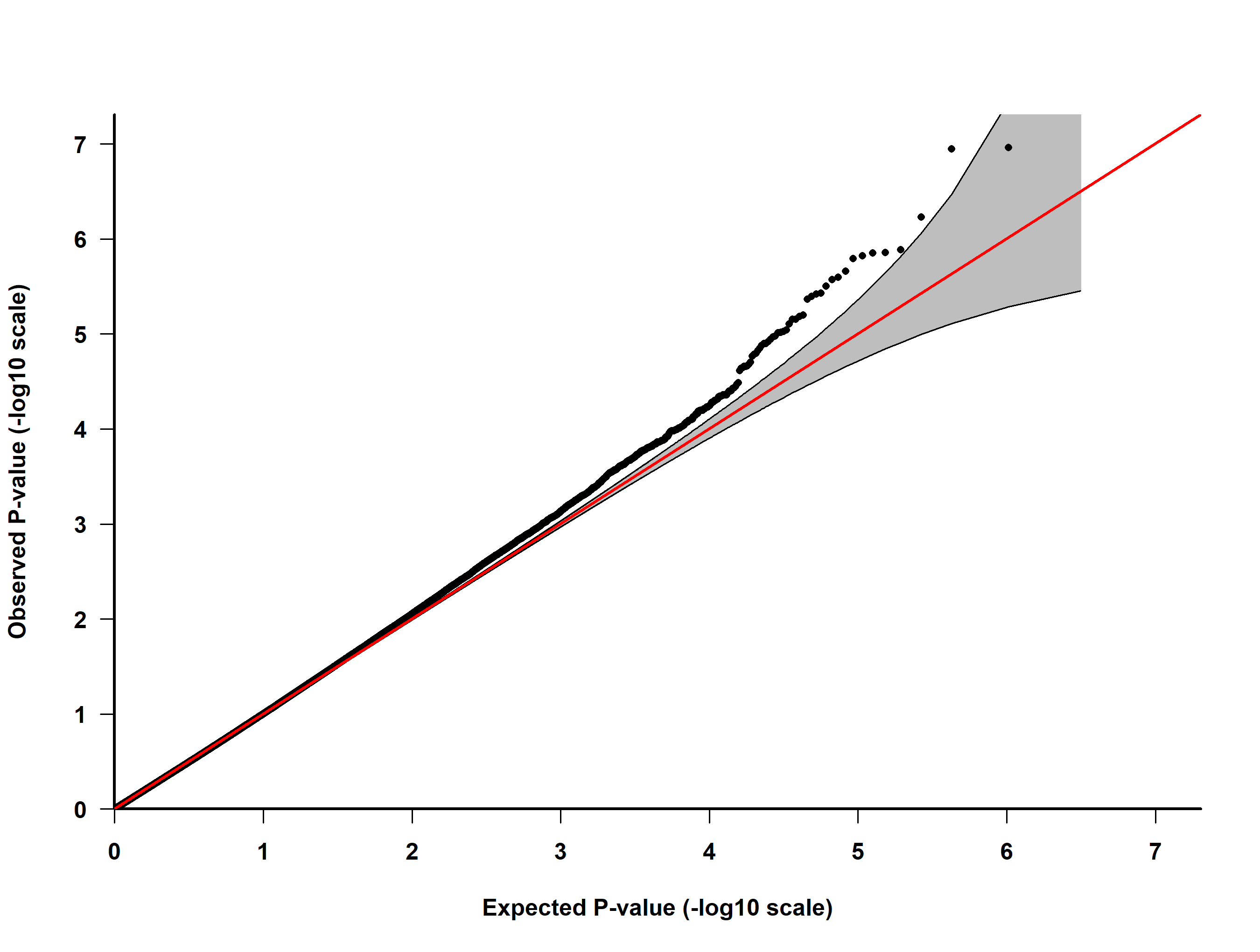

B

A

Supplementary Figure 13. QQ-plot of observed and expected *P*-values for population density in MWAS 1 (A) and MWAS 2 (B)

The straight line is where the observed *P*-values match those expected and the shaded area is the 95% confidence interval. Genomic inflation: MWAS 1 = 1.215, MWAS 2 = 0.978

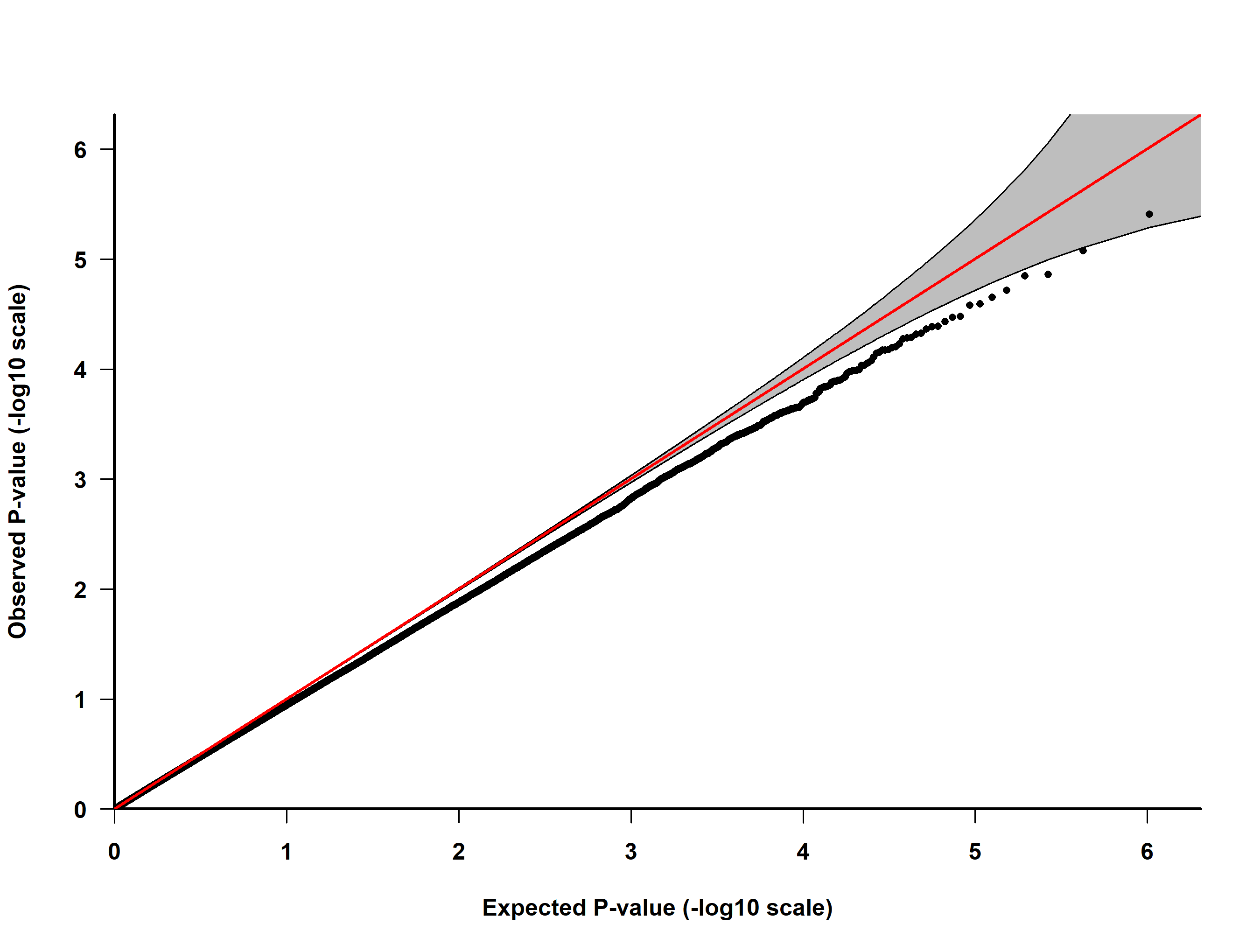

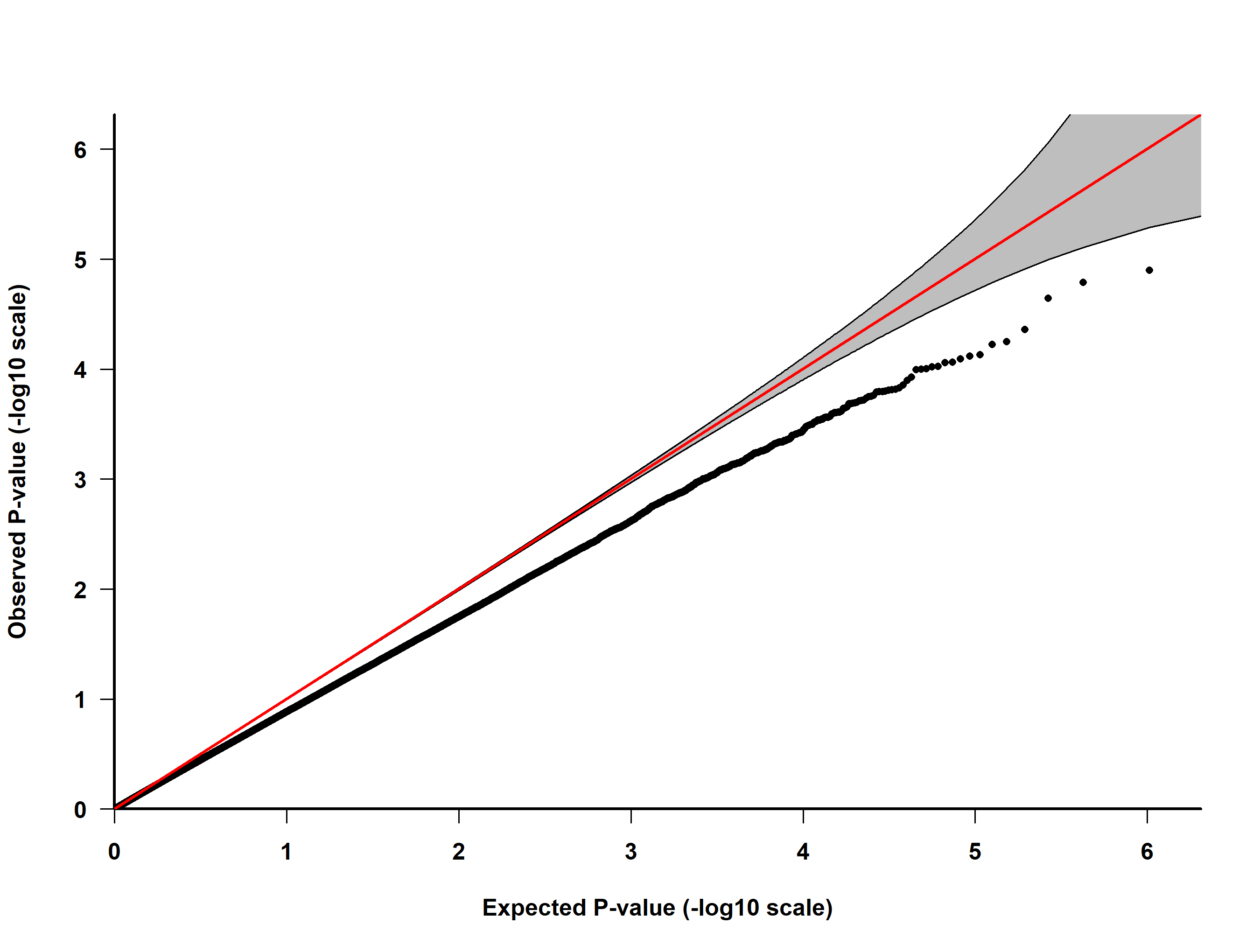

B

A

Supplementary Figure 14. QQ-plot plot of observed and expected *P*-values for urbanicity in MWAS 1 (A) and MWAS 2 (B)

The straight line is where the observed *P*-values match those expected and the shaded area is the 95% confidence interval. Genomic inflation: MWAS 1 = 0.929, MWAS 2 = 0.844

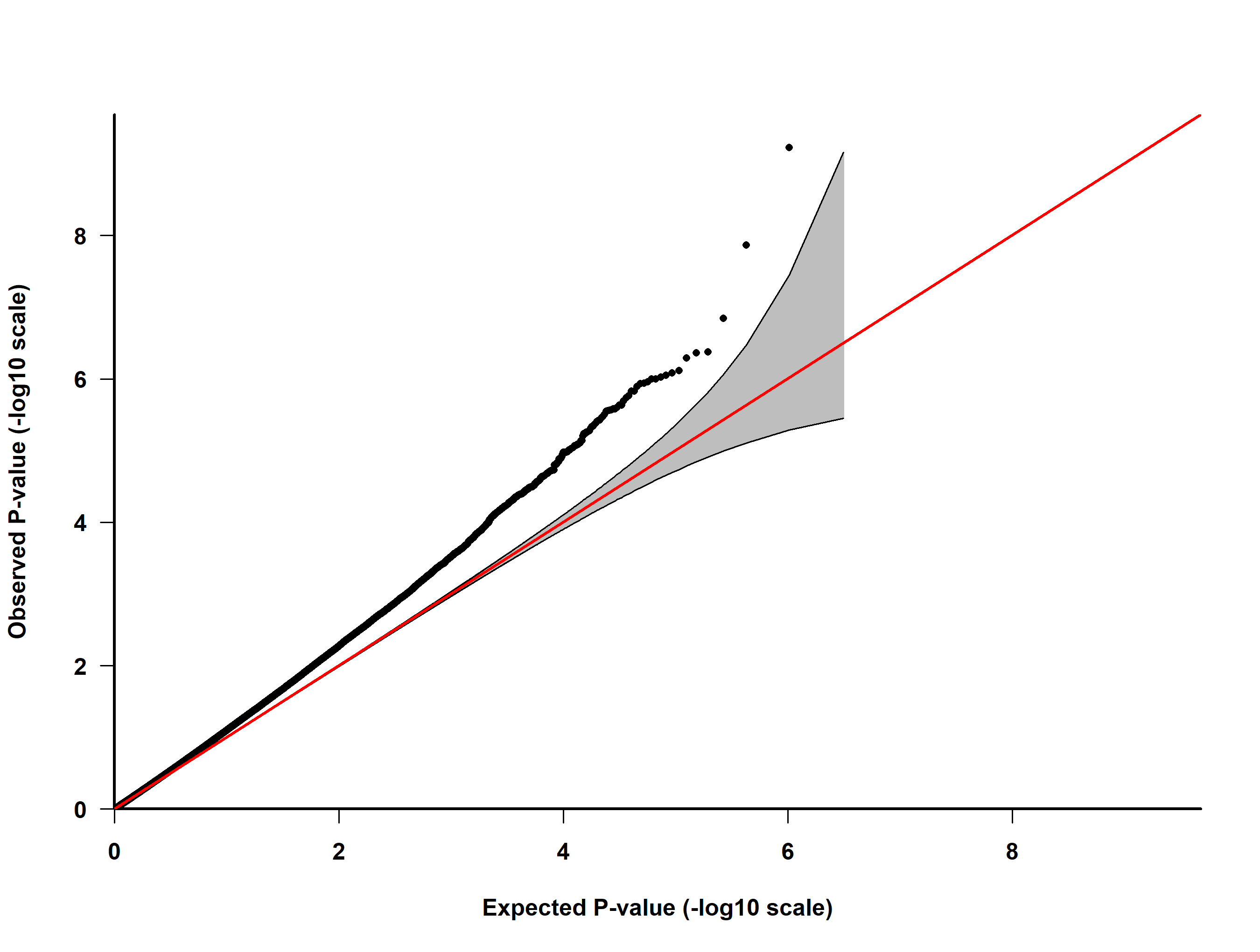

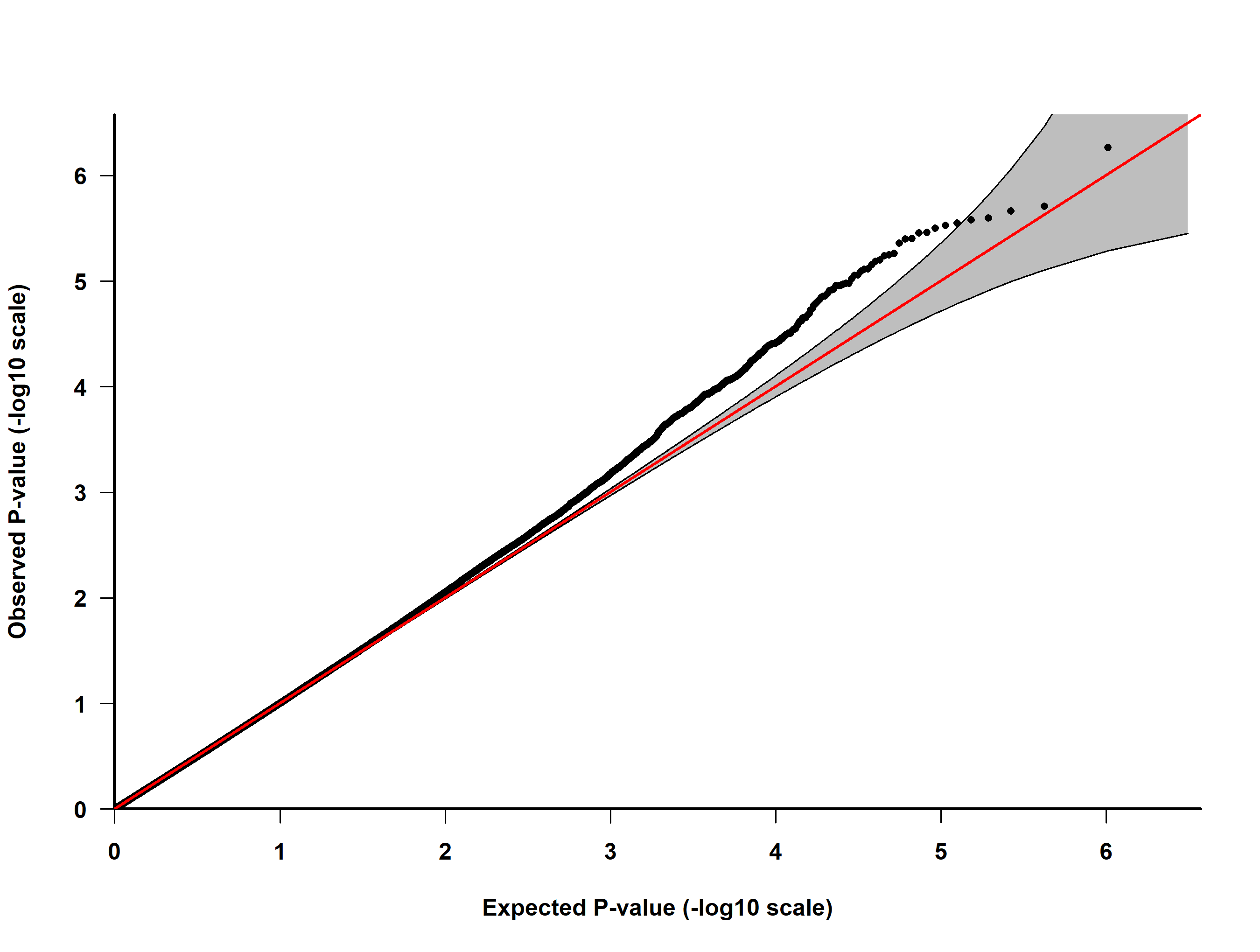

A

B

Supplementary Figure 15. QQ-plot of observed and expected *P*-values for major depressive disorder in MWAS 1 (A) and MWAS 2 (B)

The straight line is where the observed *P*-values match those expected and the shaded area is the 95% confidence interval. Genomic inflation: MWAS 1 = 1.107, MWAS 2 = 0.978

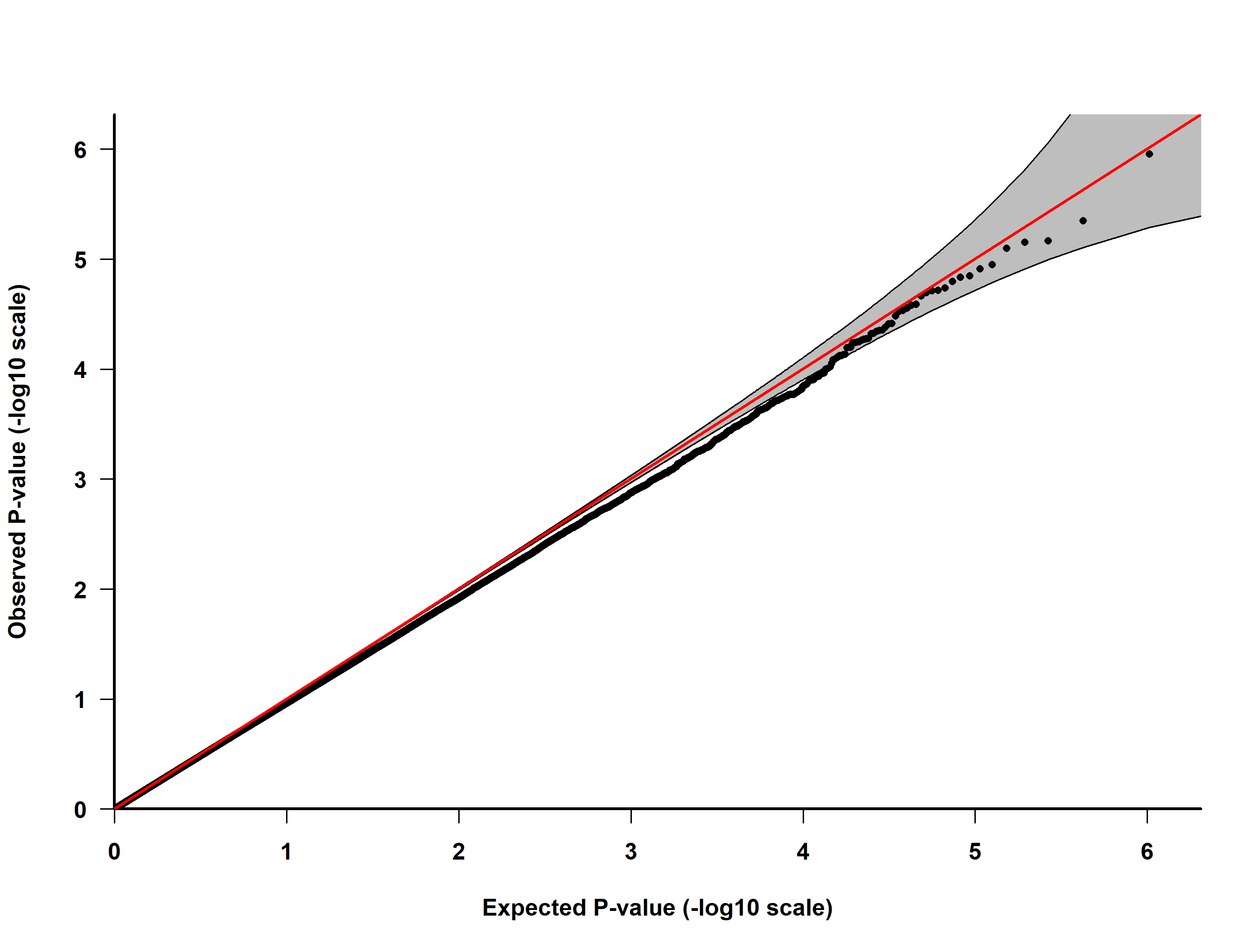

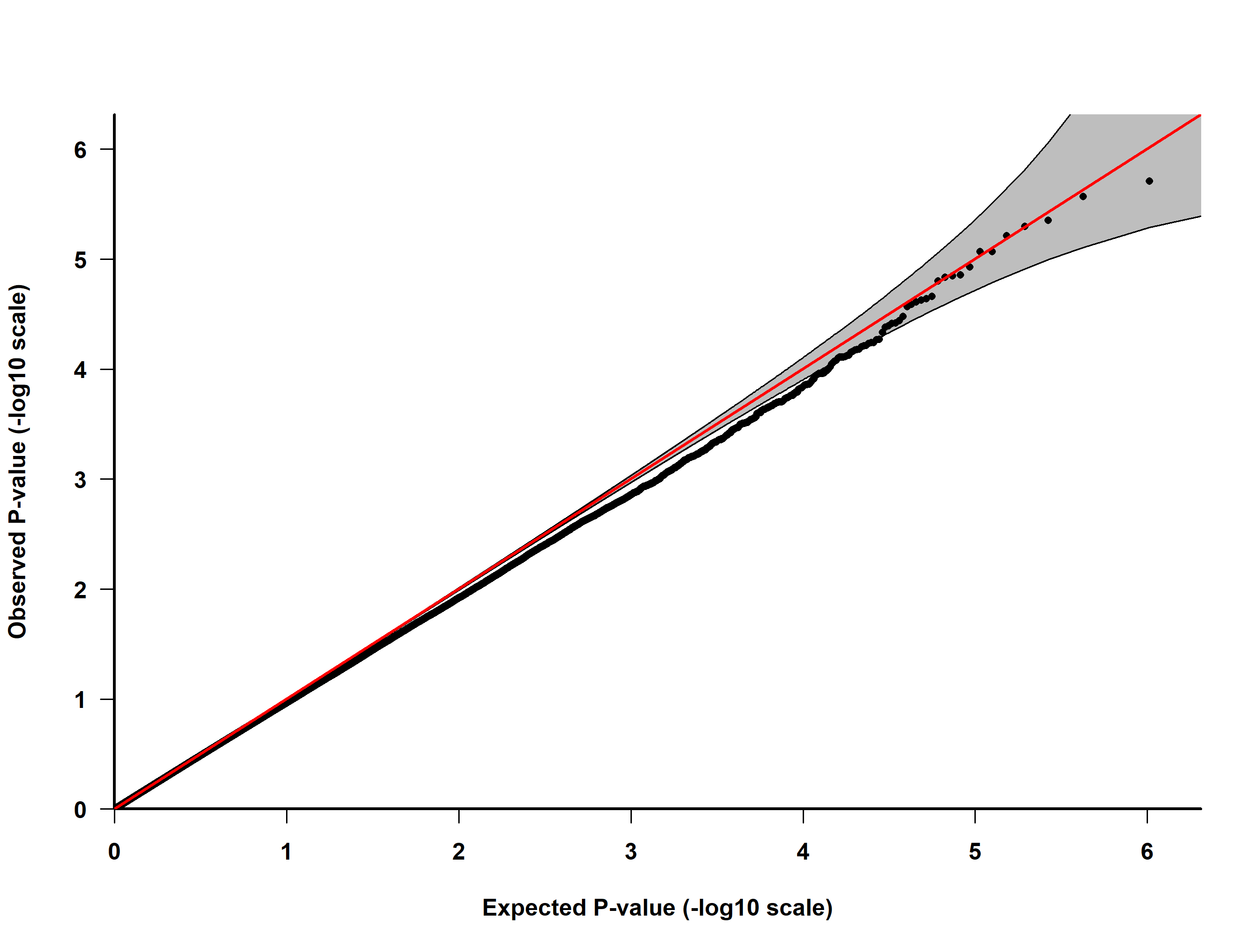

A

B

Supplementary Figure 16. QQ-plot of observed and expected *P*-values for brief resilience scale in MWAS 1 (A) and MWAS 2 (B)

The straight line is where the observed *P*-values match those expected and the shaded area is the 95% confidence interval. Genomic inflation: MWAS 1 = 0.951, MWAS 2 = 0.961

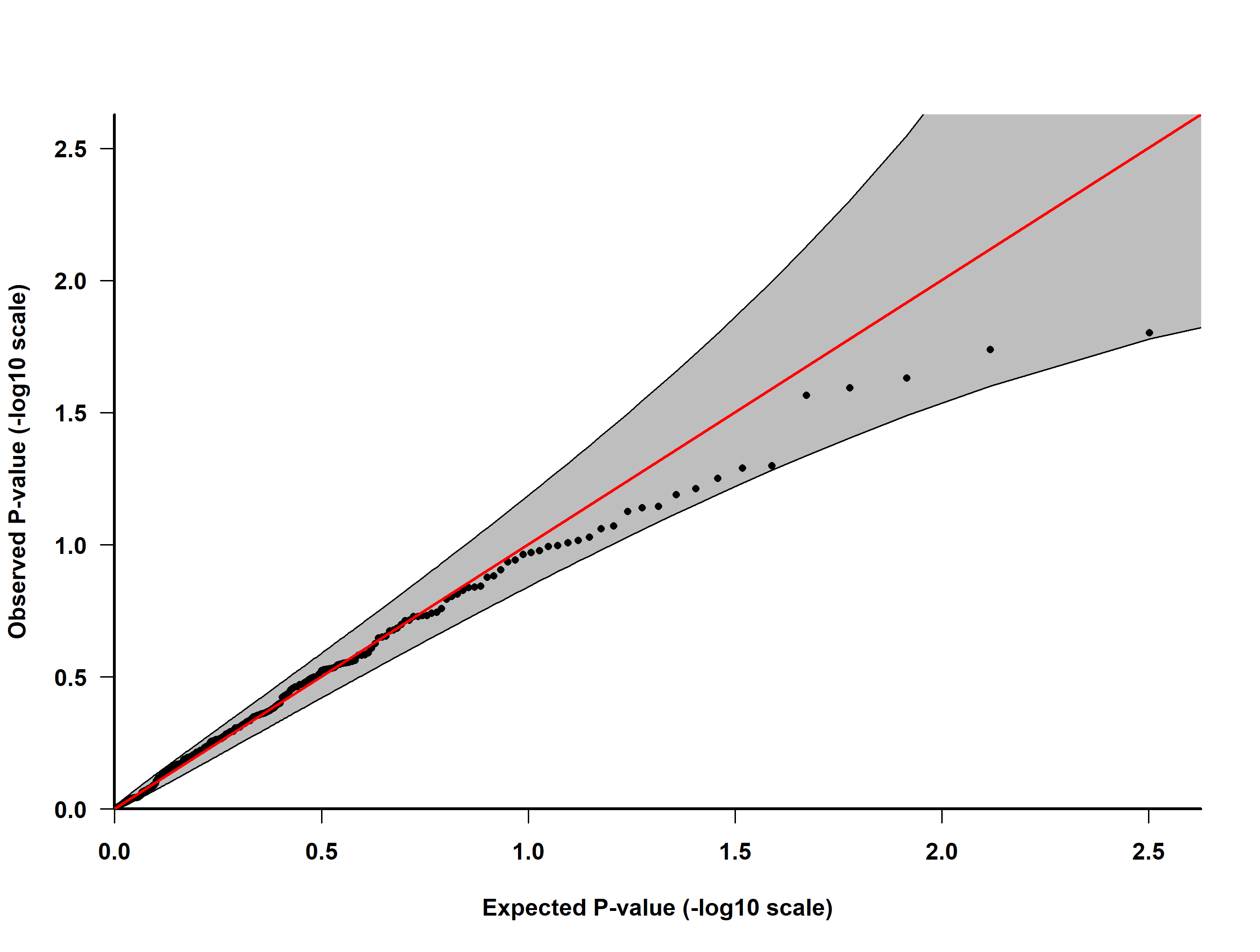

Supplementary Figure 17. QQ-plot of observed and expected *P*-values for 220 CpG sites with basophil count in the Lothian Birth Cohort

The 220 CpG sites were those associated with birth month in the Generation Scotland: Scottish Family Health Study. The straight line is where the observed *P*-values match those expected and the shaded area is the 95% confidence interval. Genomic inflation: 1.042

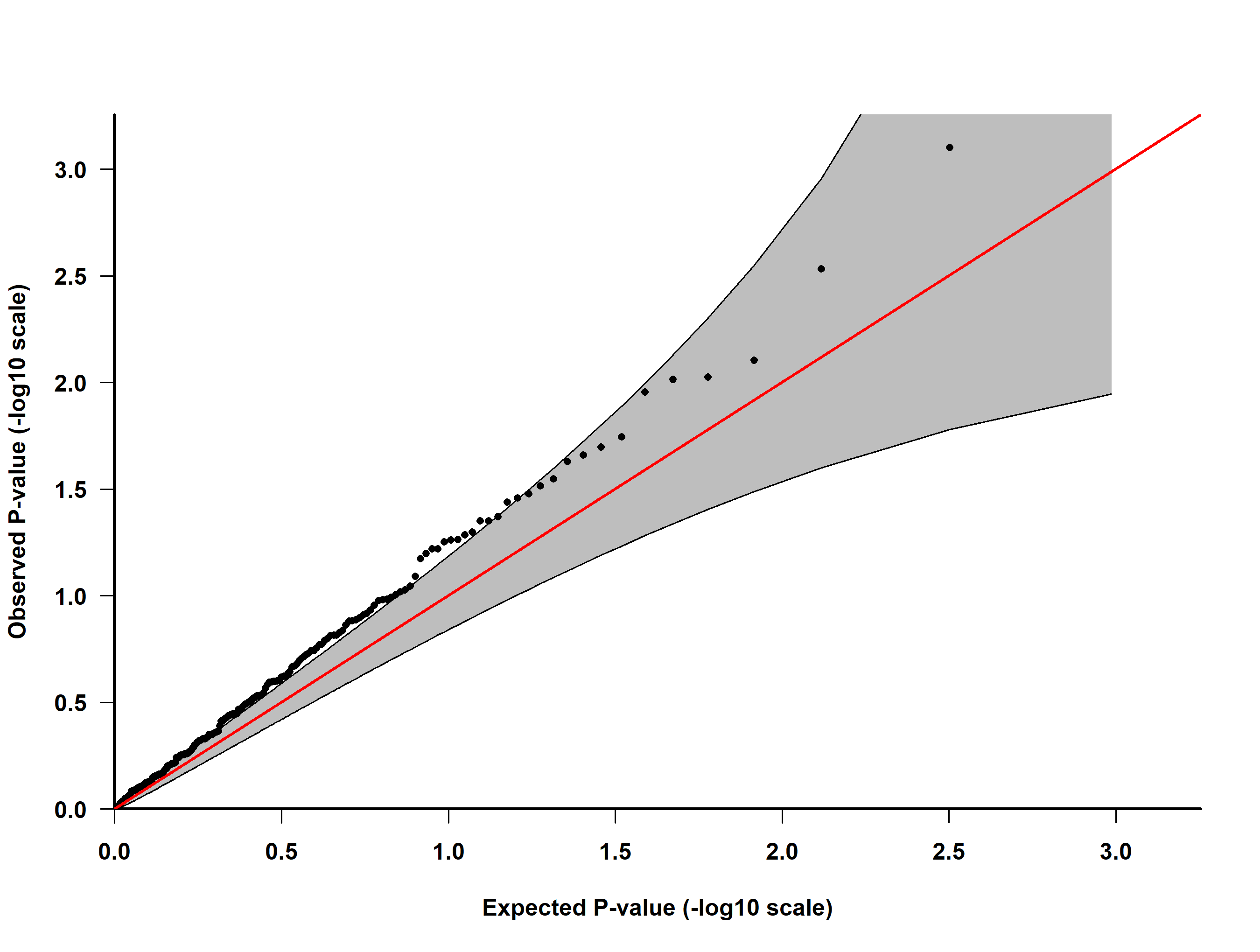

Supplementary Figure 18. QQ-plot of observed and expected *P*-values for 220 CpG sites with eosinophil count in the Lothian Birth Cohort

The 220 CpG sites were those associated with birth month in the Generation Scotland: Scottish Family Health Study. The straight line is where the observed *P*-values match those expected and the shaded area is the 95% confidence interval. Genomic inflation: 1.319

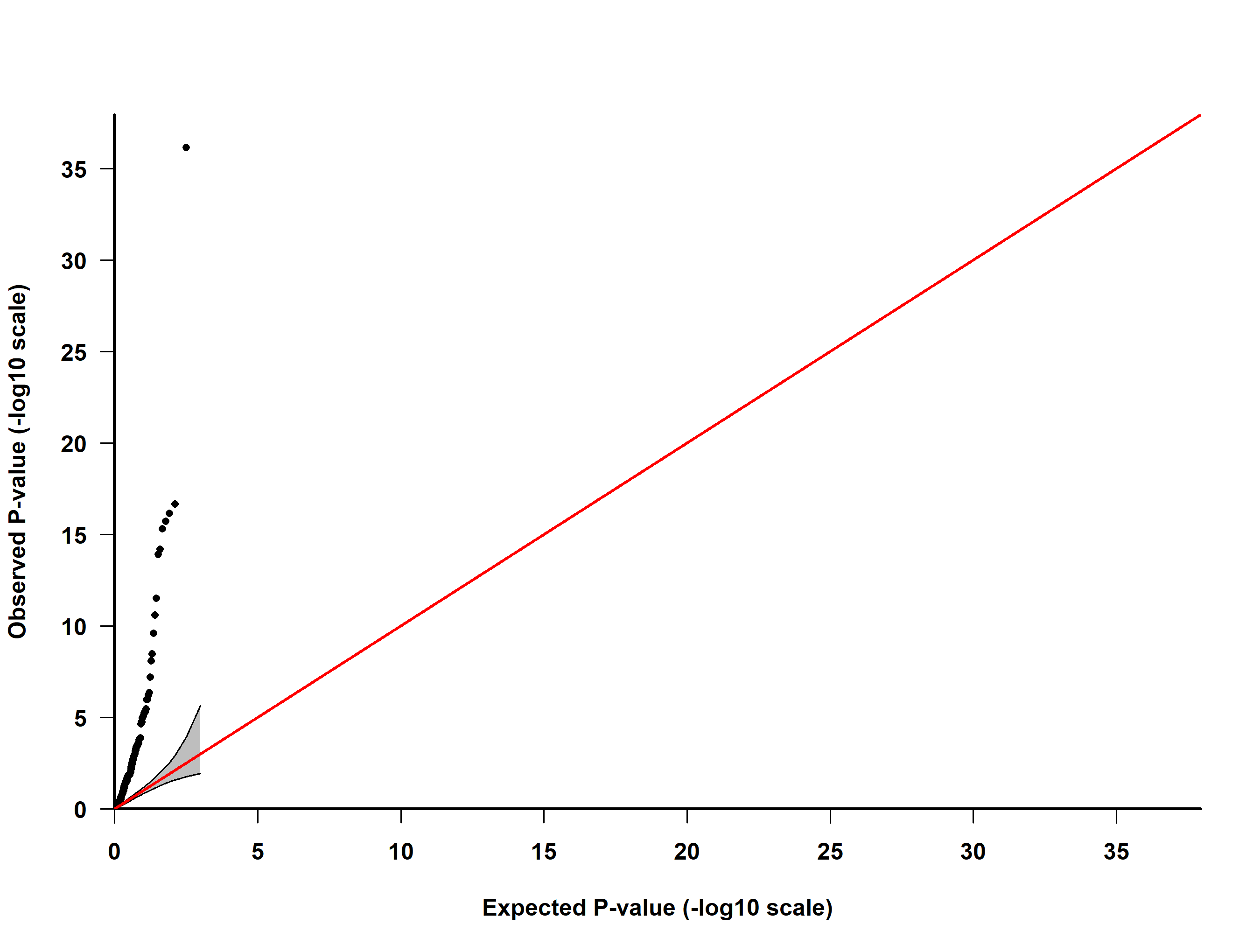

Supplementary Figure 19. QQ-plot of observed and expected *P*-values for 220 CpG sites with lymphocyte count in the Lothian Birth Cohort

The 220 CpG sites were those associated with birth month in the Generation Scotland: Scottish Family Health Study. The straight line is where the observed *P*-values match those expected and the shaded area is the 95% confidence interval. Genomic inflation: 5.841

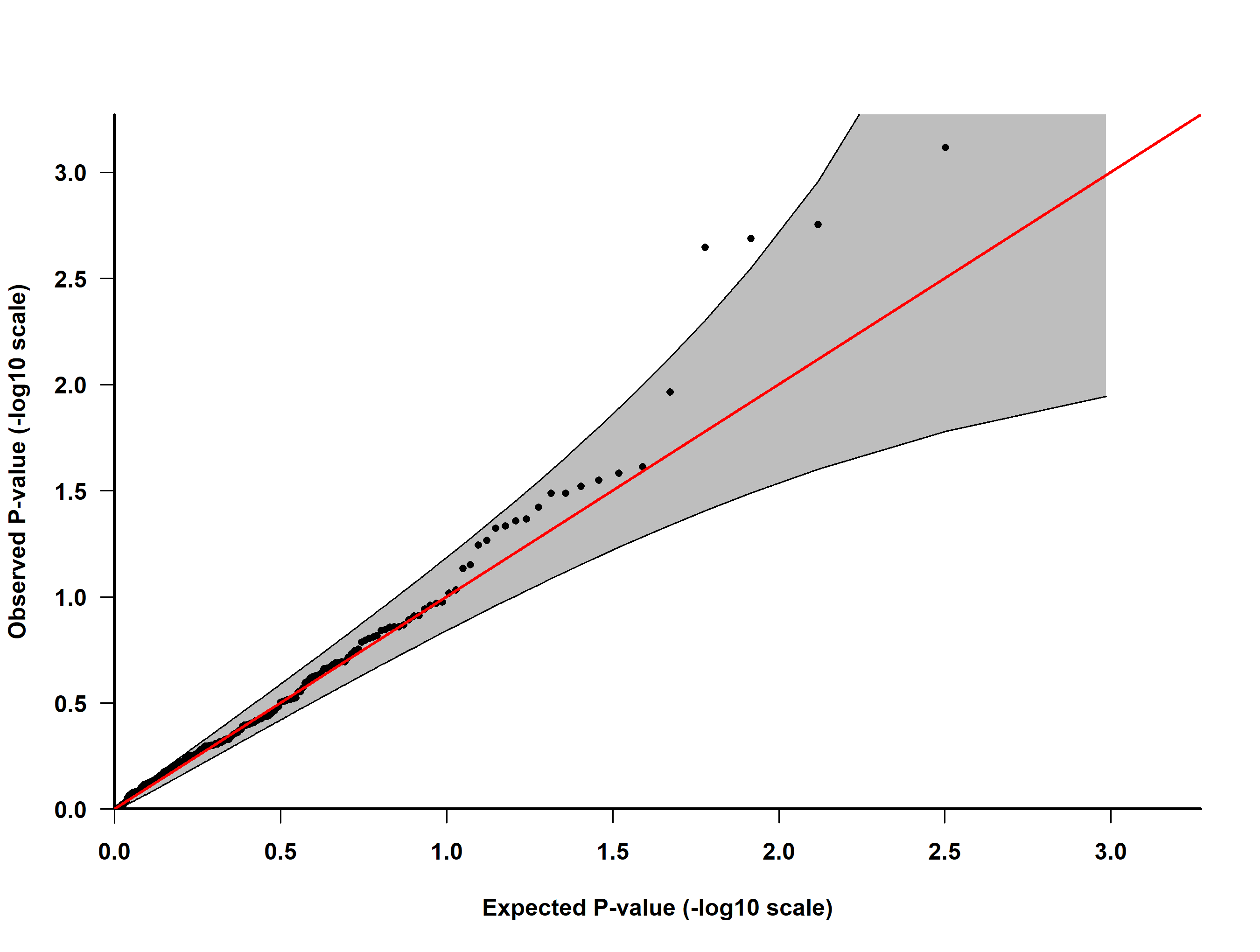

Supplementary Figure 20. QQ-plot of observed and expected *P*-values for 220 CpG sites with monocyte count in the Lothian Birth Cohort

The 220 CpG sites were those associated with birth month in the Generation Scotland: Scottish Family Health Study. The straight line is where the observed *P*-values match those expected and the shaded area is the 95% confidence interval. Genomic inflation: 1.014

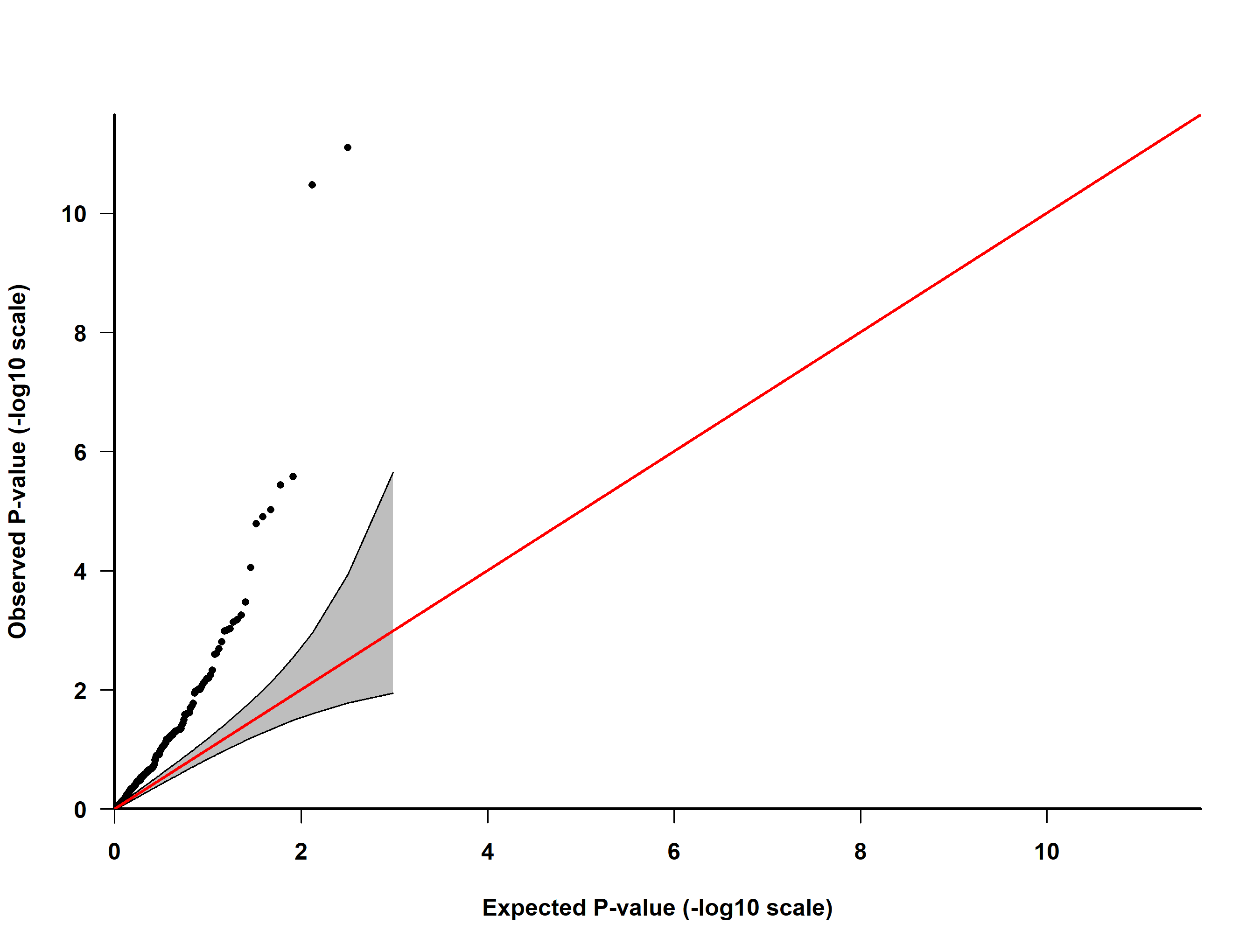

Supplementary Figure 21. QQ-plot of observed and expected *P*-values for 220 CpG sites with neutrophil count in the Lothian Birth Cohort

The 220 CpG sites were those associated with birth month in the Generation Scotland: Scottish Family Health Study. The straight line is where the observed *P*-values match those expected and the shaded area is the 95% confidence interval. Genomic inflation: 2.503
